# The words community dwelling, Spanish preferring Hispanic/Latino adults use to talk about Alzheimer’s disease and genetic testing: Implications for education and outreach

**DOI:** 10.1101/2025.08.19.25334002

**Authors:** Jamie C. Fong, Fatima I. Chavez, Karla Silos, Mirna L. Arroyo-Miranda, Gabriela Castro Castro, Mark E. Kunik, Joshua M. Shulman, Luis D. Medina

## Abstract

**INTRODUCTION:** Hispanic/Latino (H/L) adults are more likely than non-Hispanic white adults to have Alzheimer’s disease (AD), but fewer than 1 in 5 H/L adults has *APOE* ε4. H/L adults are underrepresented in AD research studies and trials, which use genetic data to stratify participants. Successful research programs representative of the entire U.S. population, including over 16 million Spanish speakers, require culturally appropriate educational materials about AD and genetic testing. We sought to learn the culturally salient words Spanish-preferring H/L adults use to talk about AD and genetic testing.

**METHODS:** Participants were community-residing and self-identified as Spanish preferring H/L adults. Fourteen individuals completed freelisting interviews, which yielded lists featuring all the words that came to participants’ minds about AD-related domains. We performed inductive thematic analysis and calculated theme frequency.

**RESULTS:** Participants were aware of AD as a memory disorder due to advancing age and genes, but were unfamiliar with AD genetic testing. Participants suggested genetic testing was more useful for diagnosis than future risk prediction. They also suggested genetic testing of individuals with intact cognition and no AD family history had limited value.

**DISCUSSION:** Findings suggested individuals are motivated by a technological imperative to participate in AD research, reflecting a responsibility to use genetic testing despite having limited knowledge about it. Interest among H/L adults in AD research could be leveraged to develop educational materials co-created by community members and researchers. Content about primary and secondary findings and the use of AD genetic results to inform a future-oriented disposition to health comprises a useful framework for AD outreach serving diverse populations.

## 1. INTRODUCTION

Alzheimer’s disease (AD) disproportionately affects older individuals. Comprising the fastest growing population in the U.S., Hispanic/Latino (H/L) individuals face accelerating rates of AD. Compared to non-Hispanic white (NHW) individuals, older H/L adults are 1.5 times more likely to have AD.^1–3^ This increased AD risk may be influenced by medical factors (e.g. genetic)^4–6^ and non-medical factors (e.g. social and structural).^7,8^

Knowledge of the genetic contribution to AD among H/L adults is incomplete. Highly penetrant, deterministic gene variants are a rare cause of autosomal dominant AD. By contrast, the ε4 allele of the *apolipoprotein E* (*APOE*) gene is a common risk factor, associated with increased AD risk among varied populations, but is present in fewer than 1 in 5 H/L adults.^9^ Current practice guidelines for the diagnostic evaluation of AD recommend genetic testing of high penetrance AD gene variants in selected scenarios, such as early-onset disease or when family history suggests autosomal dominant inheritance.^10^ Recently, *APOE* genotyping has become an important factor when considering anti-amyloid therapy due to elevated risk of ARIA,^11,12^ but it is otherwise not recommended in diagnostic evaluations. Nevertheless, *APOE* genotyping is frequently performed in research settings, where genetic data can be used with other biomarkers to stratify individuals, such as for therapeutic AD trials. However, most AD research studies and trials have included predominantly NHW adults,^13,14^ and programs serving as national resources for AD research have not yet achieved inclusion of a H/L cohort representative of the national U.S. population.^15^ Successful AD research programs benefitting the entire population, including underrepresented H/L adults, will require culturally appropriate outreach and educational materials about AD and genetic testing. In fact, the availability of Spanish language materials will be essential for community engagement, as nearly 40% of 41.4 million Spanish speakers in the U.S. have limited English proficiency.^16^ Availability of Spanish language educational materials about AD genetic testing is scant.

By contrast, Spanish language materials about cancer gene panels are more abundant. Interestingly, rates of cancer genetic test adoption among H/L adults are lower than among NHW adults, even when expertly designed Spanish language materials are used to guide informed decision making.^17,18^ This highlights the importance of developing educational materials featuring content chosen by a community of interest, rather than by experts, as salient.

In this study, we interviewed Spanish-preferring H/L adults in Houston, one of the largest and most diverse U.S. cities,^19^ using a freelisting technique to identify AD cultural concepts important to include in educational materials. A prior freelisting interview study about AD among Puerto Rican and NHW adults in the U.S. found that while AD was widely recognized as a disease of memory loss, there was variation in perceptions of other AD symptoms and causes.^20^ Among their respondents, Karlawish *et al*. noted an awareness of genes as a cause of AD, but did not explore further respondents’ perspectives about genetic testing. The results of our study provide a framework for creating community-tailored educational materials about AD and genetic testing for research and clinical care.

## 2. METHODS

### 2.1 Eligibility criteria

Adults (ɥ18 years) in Houston, Texas, who self-identified as Spanish-preferring (comfortable conversing entirely in Spanish) and H/L were screened. The Baylor College of Medicine Institutional Review Board approved the study. Participants gave verbal informed consent and received gift cards.

### 2.2 Recruitment and enrollment

We used a community-engaged approach—well demonstrated to foster trust and benefit between academic and community partners^21–23^—to access recruitment sources and build capacity in the Houston-area H/L community. Flyers were distributed through partners at H/L health and cultural fairs, immigration events, and adult learning classrooms. Recruitment also used social media, the local Alzheimer’s Association chapter, H/L-serving health clinics, and nonprofits serving older adults and English-as-second-language learners. Snowball sampling allowed participants to refer others from their networks. Eligible individuals contacted the team directly or via an online survey. Spanish-English bilingual-bicultural H/L coordinators followed up by phone or email to describe the study, confirm eligibility, and conduct interviews. Thirty individuals contacted the team; 7 did not respond to follow-up, 6 were ineligible, 2 declined, and 1 withdrew due to discomfort completing the interview in Spanish. Fourteen completed interviews (Figure 1).

**Figure 1.**
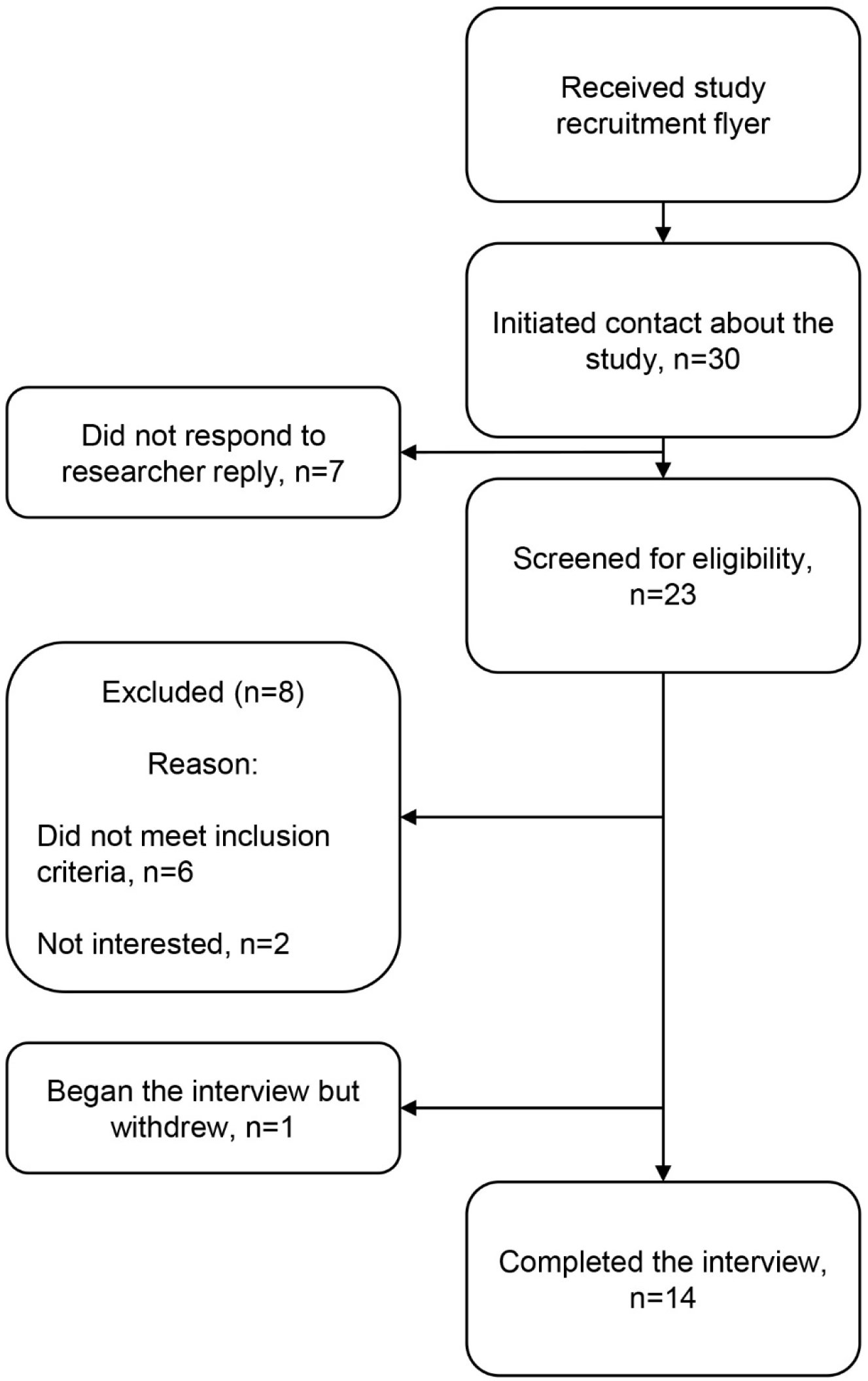
Recruitment of study population

### 2.3 Procedures

From May—September 2021, bilingual-bicultural H/L coordinators (F.C., G.C.) conducted recorded phone or videoconference interviews entirely in Spanish, comprising 9 word-cluster prompts (Supplemental Table 1) and 4 vignettes with a prompt (Supplemental Table 2). Prompts focused on words for AD, dementia, genes, and reasons for genetic testing. Vignettes described disclosure of AD genetic testing results to a fictional H/L man whose demographics, cognition, function, and genetic result varied. The vignette prompt addressed his reaction to the result. Participants also answered demographic, cultural identity, subjective understanding, and research knowledge questions. Validated measures assessed acculturation,^24^ cultural values,^25^ and subjective health literacy.^26,27^ We used established freelisting methods^28^ with supplementary techniques^29^ to maximize output.

Freelisting generates word lists reflecting culturally salient knowledge, attitudes, and discourse about a topic.^28,30^ In this study, prompts and vignettes were developed by experts in AD genetic counseling, neurology, cultural neuropsychology, and H/L cognitive aging outreach. Interviewers asked participants to list all words evoked by the prompts or the fictional man’s reaction.

### 2.4 Data analysis

Interview recordings were transcribed in Spanish and translated into English by a professional service (Landmark Associates, Inc). Transcripts in both languages facilitated team discussion, which included non-Hispanic/non-Spanish speakers and bilingual-bicultural H/L members.

Although freelist analysis^28^ was initially planned, participants frequently provided narrative rather than single-word responses. We therefore organized responses into word-phrase lists and applied inductive thematic analysis. Two bilingual-bicultural H/L coordinators (F.C., K.S.) coded transcripts in Spanish, adhering to participants’ language and meaning without preconceived assumptions. After jointly coding an initial subset and resolving differences, coders analyzed remaining transcripts independently, meeting regularly to ensure consistency. Coding results were translated into English for further coding by the full team. The same process was applied to vignette responses. Instead of summarizing word frequency, we summarized theme frequency as the number of times a theme appeared divided by the total number of unique responses.

## 3. RESULTS

### 3.1 Participants

As shown in Table 1, respondents ranged in age from 19 to 73 years (mean=49.3±16.8). All respondents identified as Mexican, Chicano, or Mexican/Chicano American, and 11 of 14 were born in Mexico.

**Table 1.**
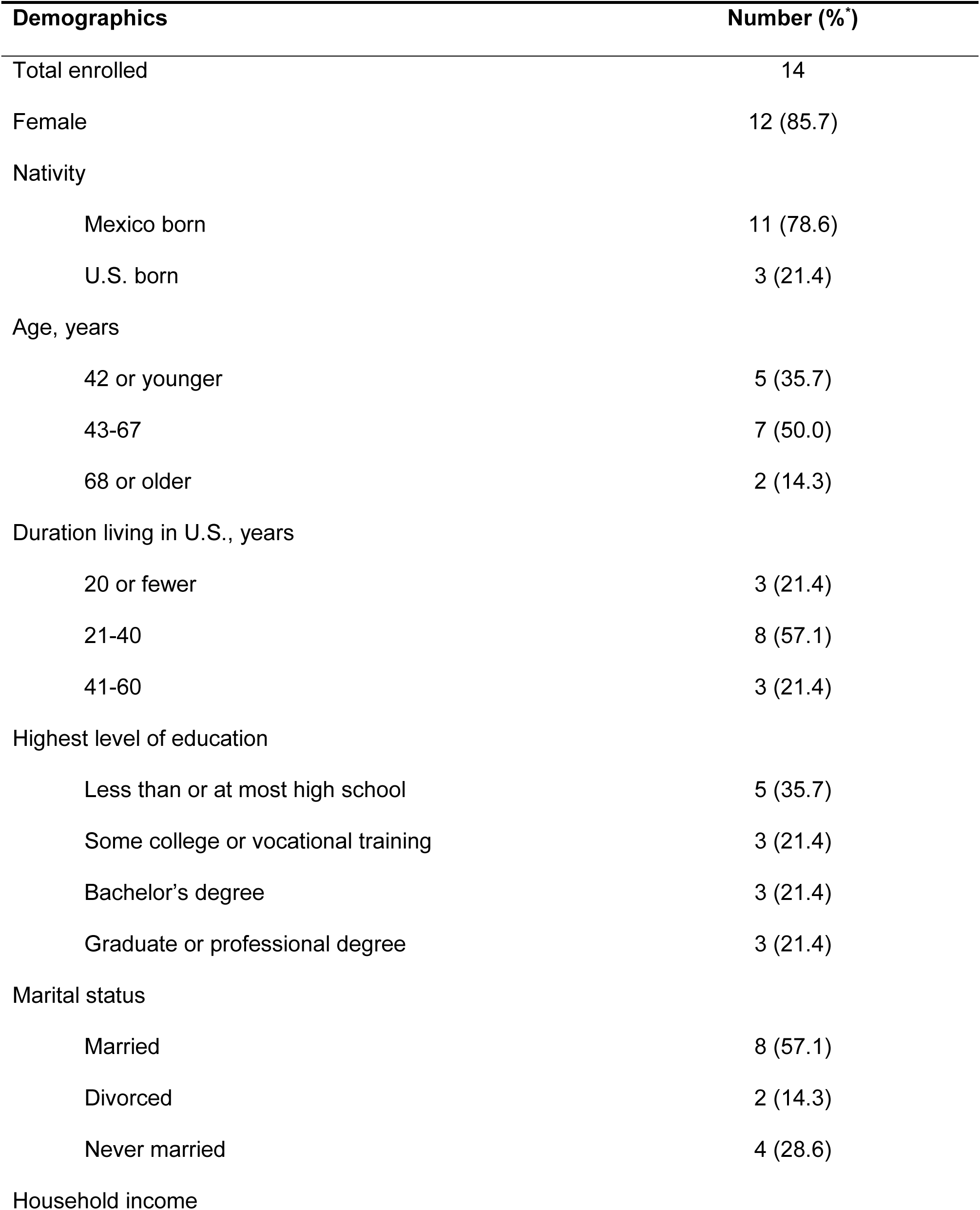
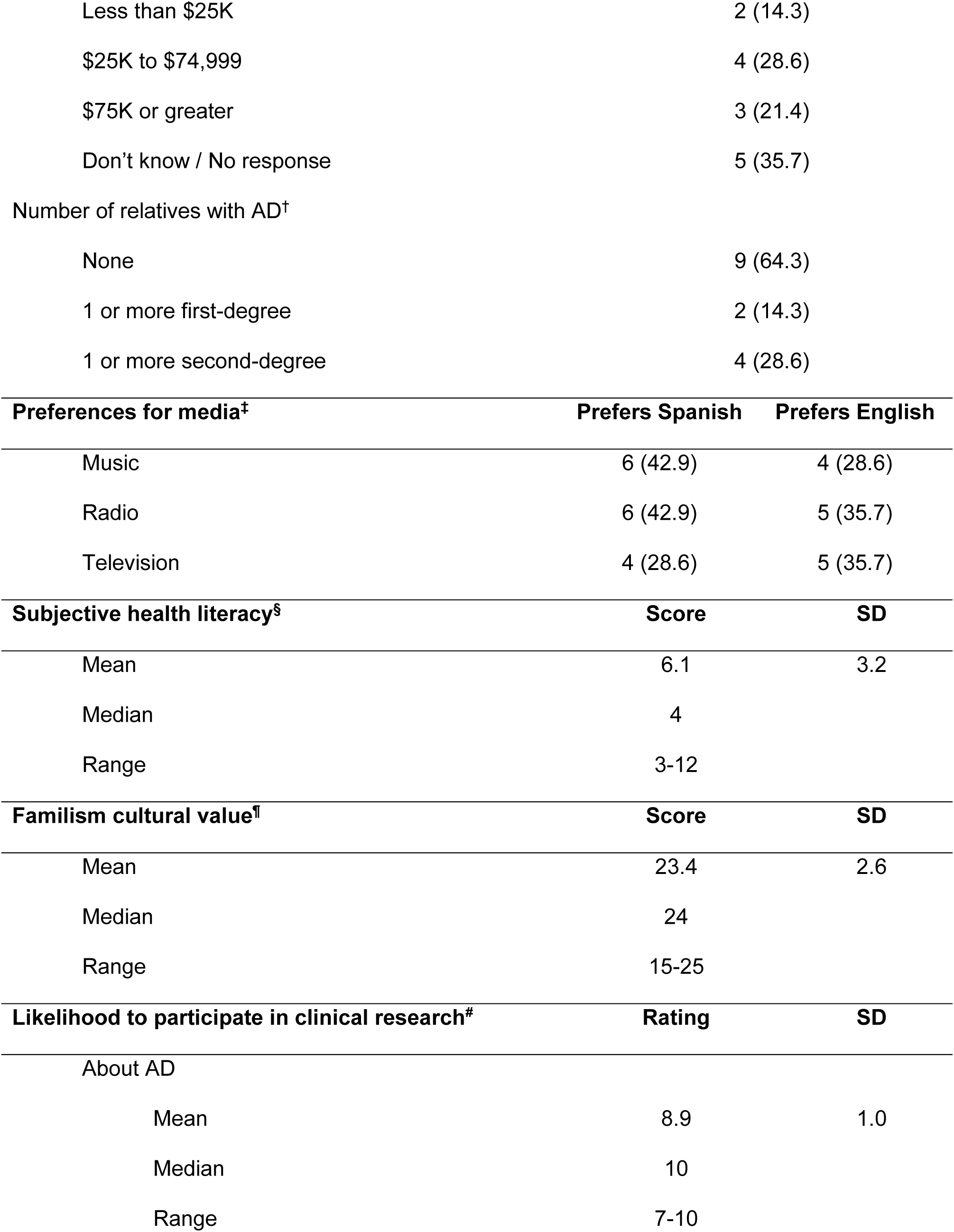
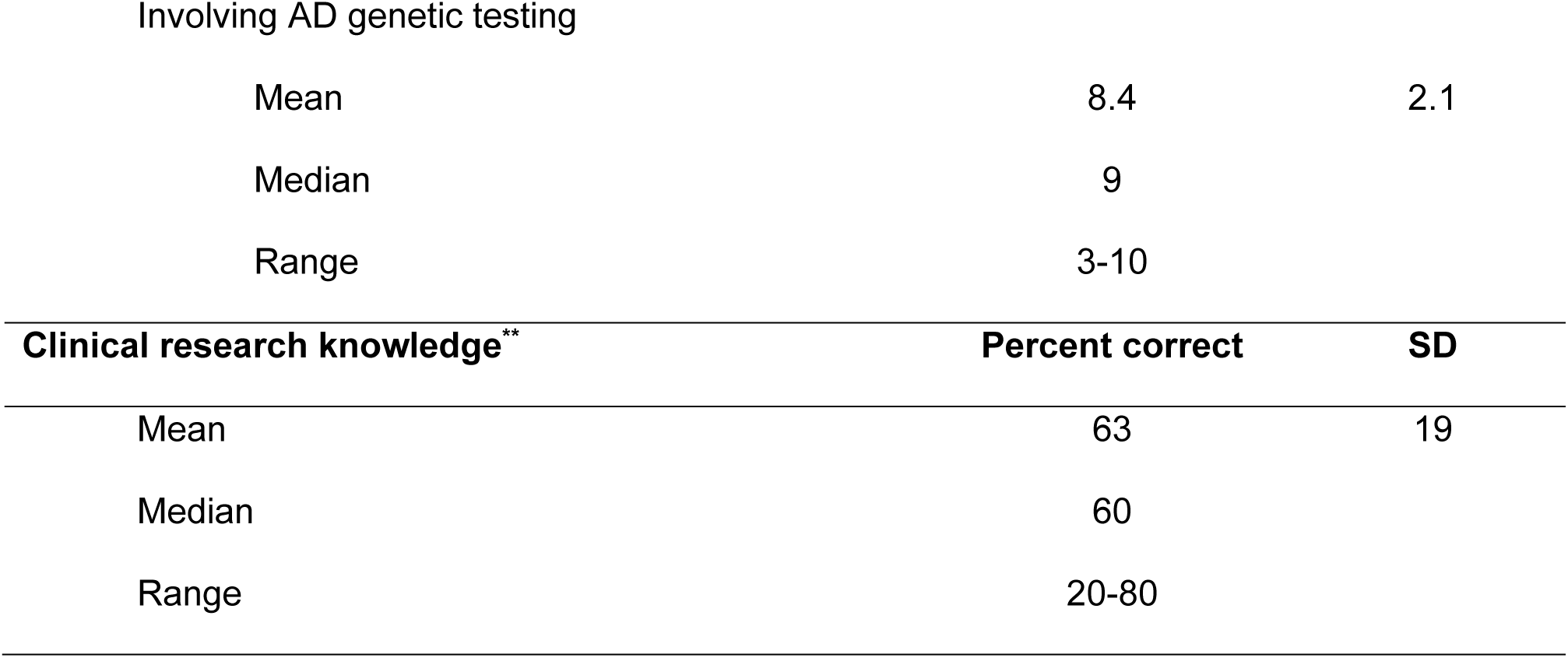
Respondent characteristics. Abbreviations: AD=Alzheimer’s disease, SD=standard deviation. *Percentages may not add up to 100% due to rounding. ^†^Percentages may not add up to 100% due some respondents having both first- and second-degree relatives with AD. ^‡^Measure of preferences for media. Three statements assessed preference for language of 3 forms of media—music, radio, and television—each with choice of “prefer Spanish”, “prefer English”, or “no preference”. Proportion of “no preference” responses is not shown, but is the difference between total enrolled and sum of proportion of other preferences. ^§^3-Brief Screening Questionnaire.^26,27^ Responses to 3 individual questions were based on a rating scale from 1 to 5. Scores ranged from 3-15, with larger scores reflecting worse self-reported health literacy. ^¶^Pan-Hispanic Familism Scale.^25^ Responses to 5 individual questions were based on a rating scale of 1-5. Scores ranged from 5-25, with larger scores reflecting greater alignment with H/L cultural value of *familismo*. ^#^Rating scale, with 1=not at all, 10=extremely likely. **Measure of clinical research knowledge. Five true-false statements assessed knowledge of clinical research, including its distinction from medical care, its voluntary nature, and other features of the informed consent process. Percent correct scores ranged from 0-100%.

Respondents resided in the U.S. for a range of 15 to 68 years (mean=44.0±17.9). Over one third had at most a high school education. Nine of 14 participants had no family history of AD. On average, participants had higher engagement with Spanish than English (Bidimensional Acculturation Scale, mean Δ= -2.6±4.1, range= -9 to 6; Table 2). In addition, language preferences for music, radio, and television varied among participants, with responses distributed across a preference for Spanish, English, or no preference. Participants’ subjective understanding of genetics and AD varied widely, though the mean rating was similar for both, falling near the midpoint of the scale that spanned from “not at all” to “extremely well” understood (Figure 2). Although participants reported a high level of understanding of clinical research (mean=8.1±2.0; Figure 2), their performance on an objective measure assessing knowledge of informed consent topics suggested a more limited grasp of key concepts (mean score=64±19%; Table 1). Participants reported high interest in clinical research involving AD genetic testing (mean=8.4±2.1; Table 1).

**Figure 2.**
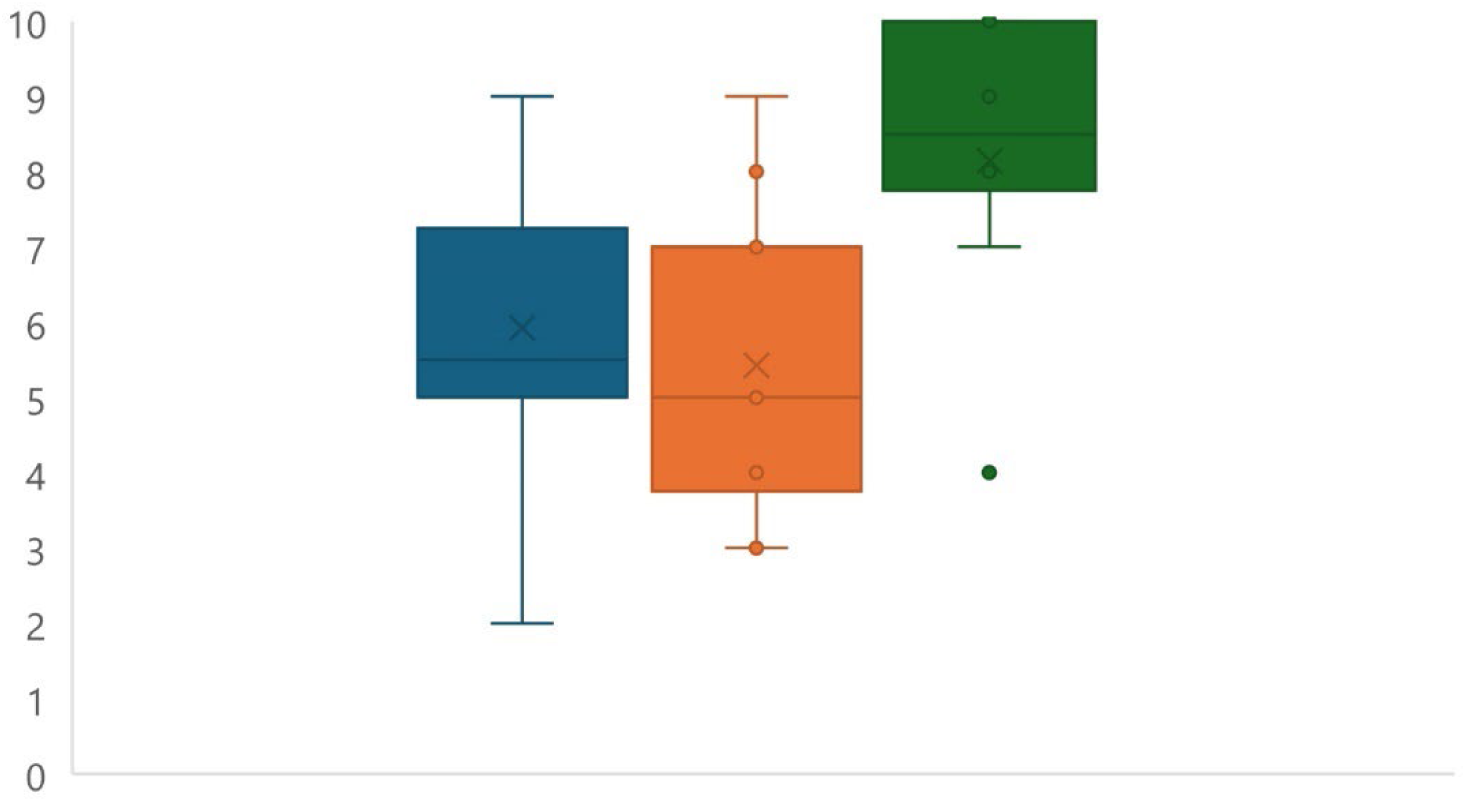
Subjective understanding. Responses to rating scales measured self-reported understanding of genetics, Alzheimer’s disease (AD), and clinical research. Each rating ranged from 1=not at all to 10=extremely well understood. Higher ratings reflected greater perceived understanding. Blue=genetics (mean=5.9±1.8, median=5.5, range=2-9), orange=AD (mean=5.4±2.0, median=5, range 3-9), green=clinical research (mean=8.1±2.0, median=8.5, range=4-10).

**Table 2.** Bidimensional Acculturation Scale for Hispanics.^24^ For each language subscale (English, Spanish), responses to 3 individual questions were based on a rating scale of 1-5. Scores ranged from 3-12, with larger scores reflecting high engagement with the subscale language. Delta (Δ) represents the difference between English and Spanish subscale scores, with a greater negative value associated with greater Spanish than English language engagement, and Δ=0 suggesting bicultural, bilingual engagement.

| | English | | | Spanish | | | Delta ($\Delta$) | | | |
| --- | --- | --- | --- | --- | --- | --- | --- | --- | --- | --- |
|  | Mean | SD | Range | Mean | SD | Range | Mean | SD | Min | Max |
| Subscale score | 8.1 | 2.9 | 3-12 | 10.8 | 1.7 | 6-12 | -2.6 | 4.1 | -9 | 6 |

### 3.2 Participants have limited familiarity with AD genetic testing

Interviews began by prompting participants with words and phrases about AD, dementia; genes; and reasons for genetic testing, and we analyzed responses to identify dominant themes. Detailed results are shown in Table 3, and we focus here on excerpts of potential importance. Respondents believed AD was a serious illness. When asked about “illnesses worse than AD,” only “cancer” *(cancer)* and “mental illness” *(enfermedades mentales)* were cited more frequently (20% and 11%). Across responses to all but two prompts, the theme “don’t know” *(no sé)* emerged at a frequency of 2-7%, reflecting limited familiarity among a minority of respondents. The theme “don’t know” *(no sé)* comprised none of the responses to the prompts about “genes” and “illnesses worse than AD,” reflecting awareness of these concepts. Prompts about “reasons to have an AD genetic test” and “more useful tests than an AD genetic test” yielded the greatest proportions of “don’t know” *(no sé)* (7% and 6%).

**Table 3.**
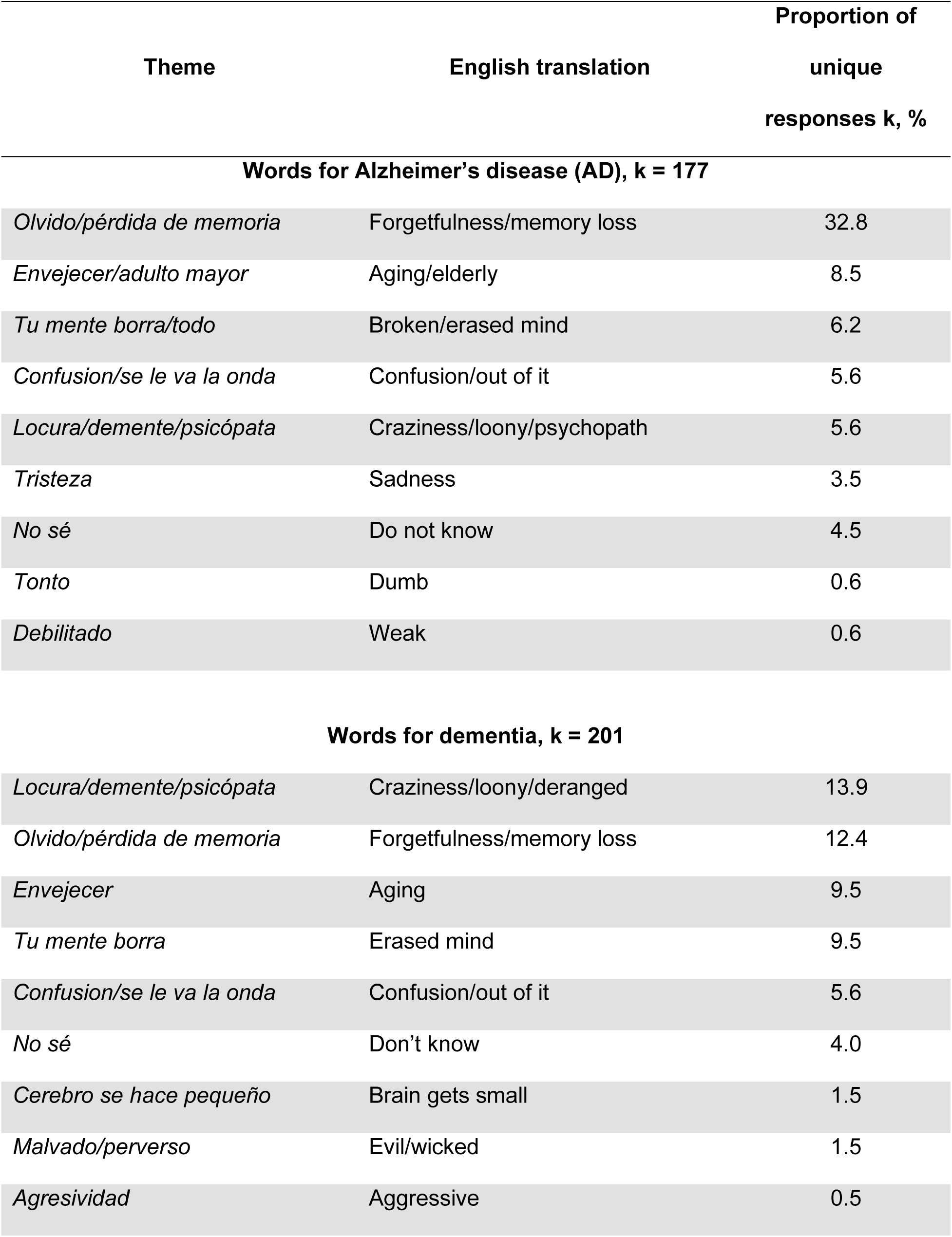
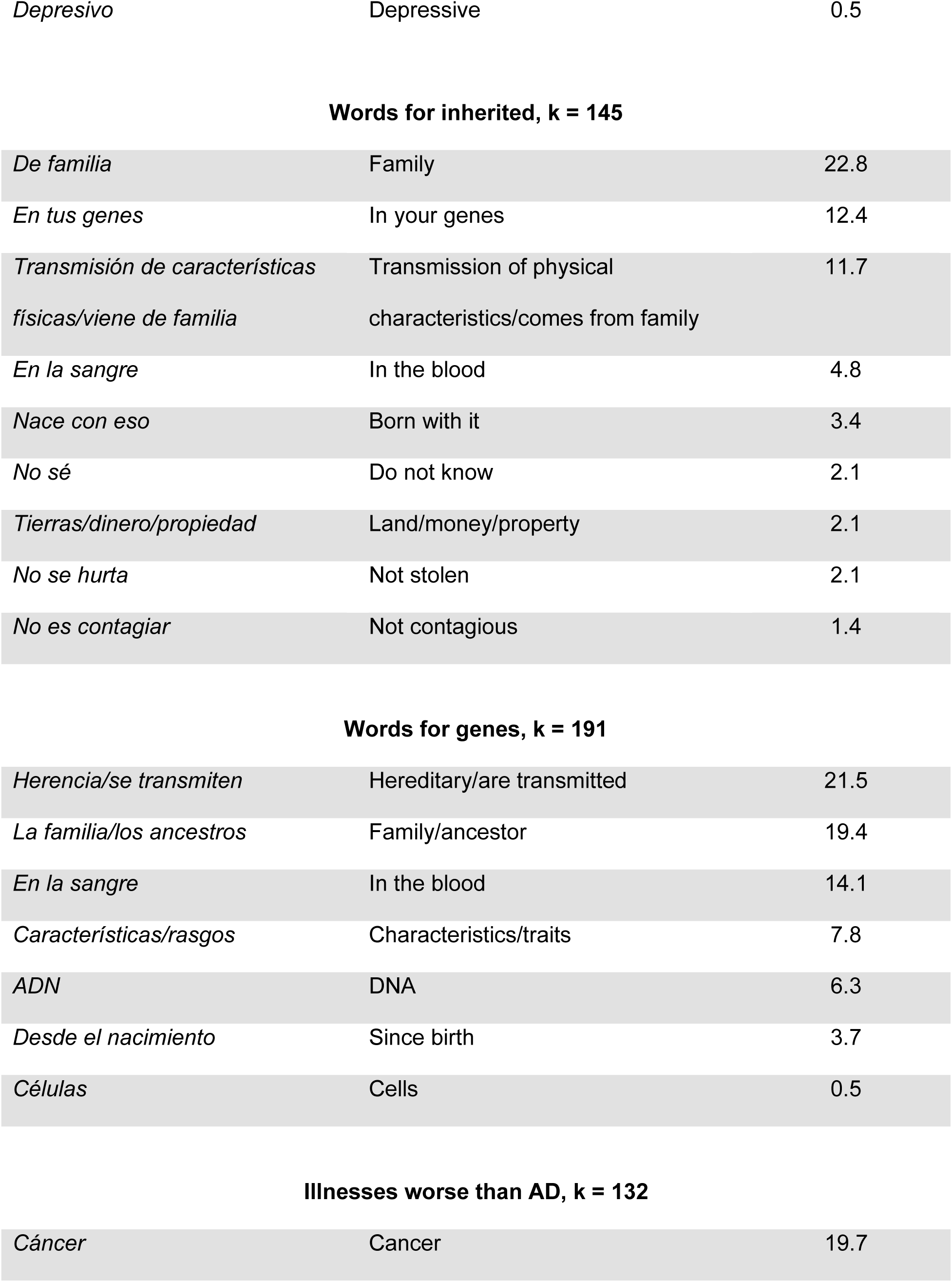
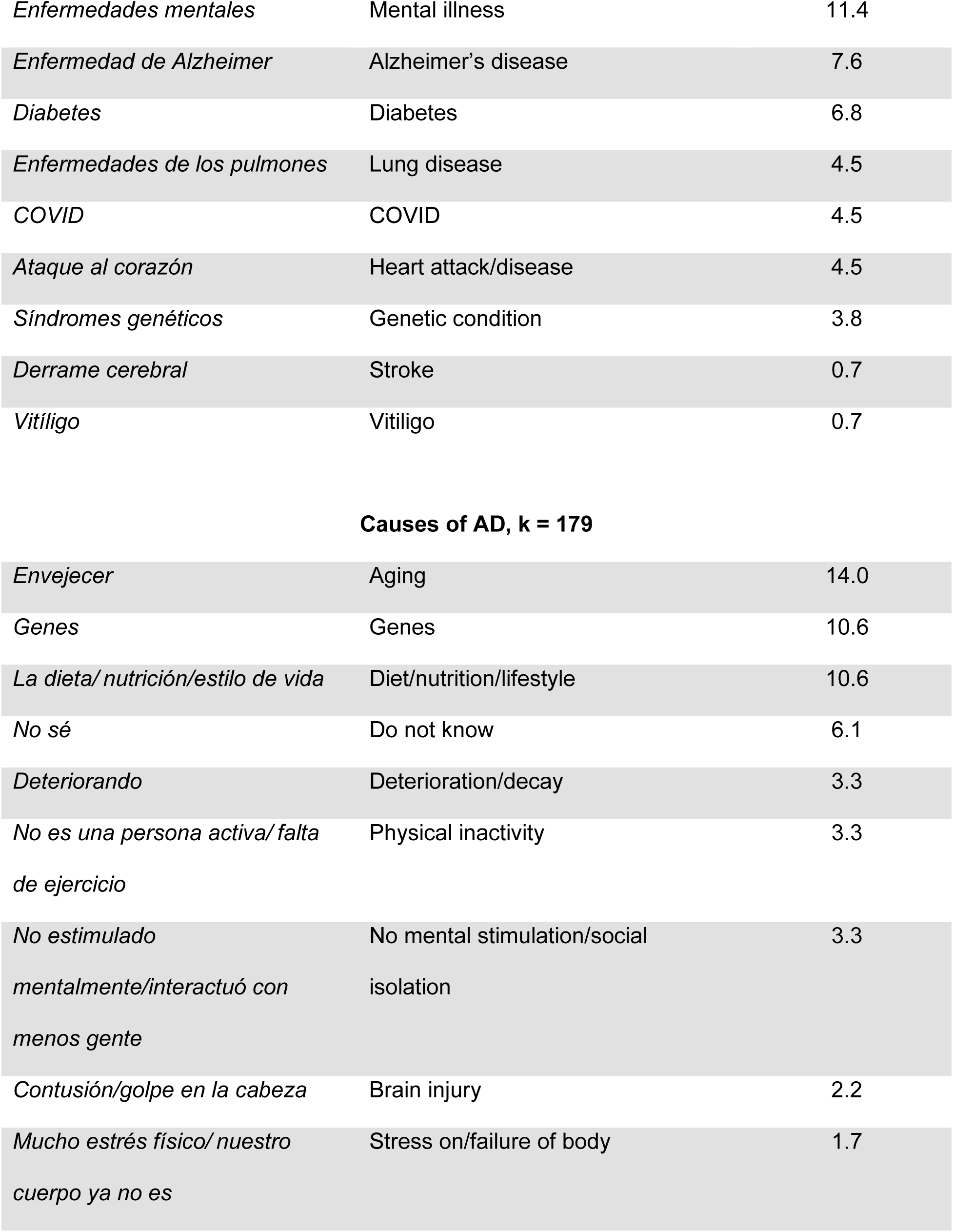
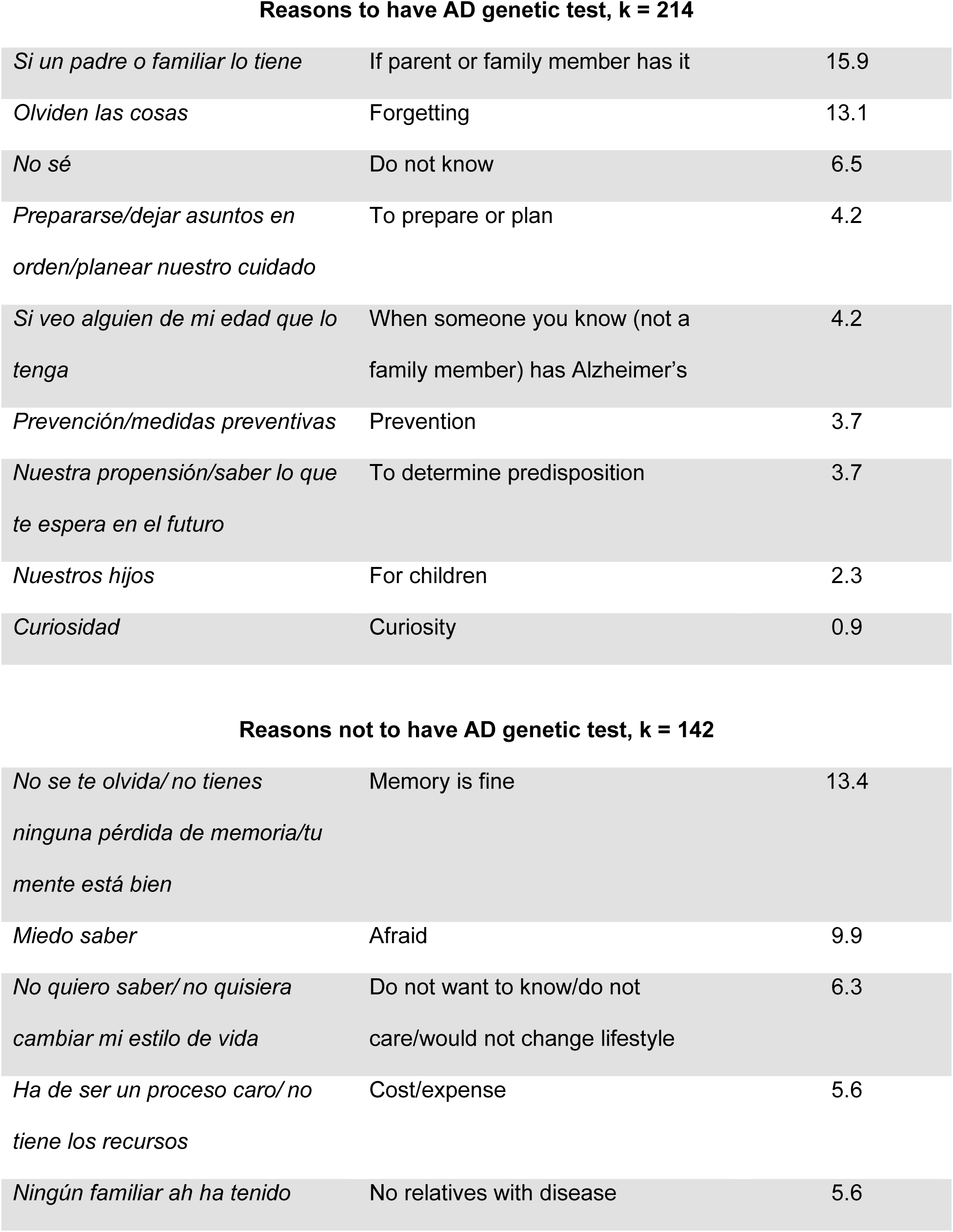
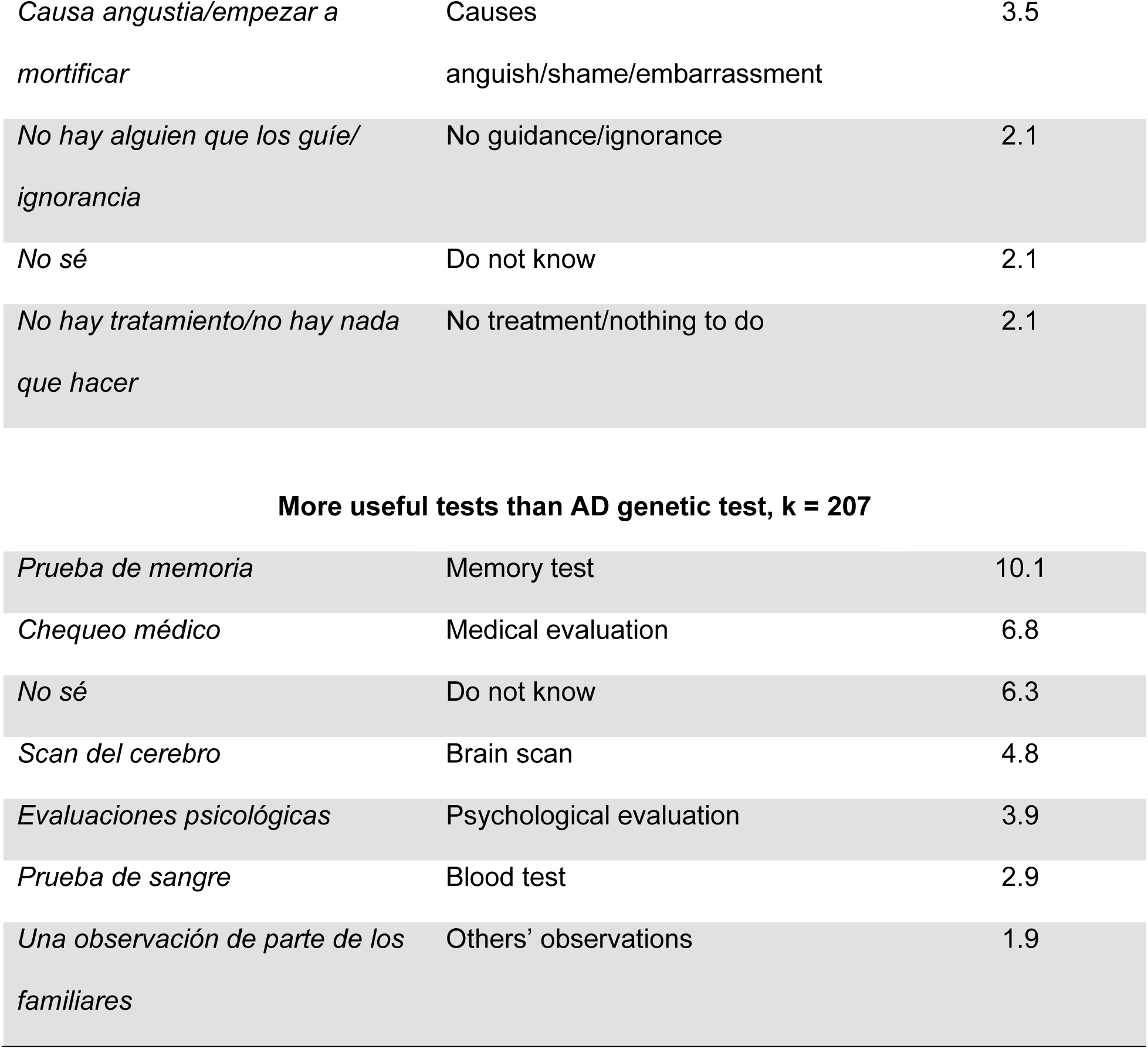
Results of word cluster prompts.

| Theme | English translation | Proportion of unique responses k, % |
| --- | --- | --- |
| **Words for Alzheimer's disease (AD), k = 177** |  |  |
| *Olvido/pérdida de memoria* | Forgetfulness/memory loss | 32.8 |
| *Envejecer/adulto mayor* | Aging/elderly | 8.5 |
| *Tu mente borra/todo* | Broken/erased mind | 6.2 |
| *Confusion/se le va la onda* | Confusion/out of it | 5.6 |
| *Locura/demente/psicópata* | Craziness/loony/psychopath | 5.6 |
| *Tristeza* | Sadness | 3.5 |
| *No sé* | Do not know | 4.5 |
| *Tonto* | Dumb | 0.6 |
| *Debilitado* | Weak | 0.6 |
| **Words for dementia, k = 201** |  |  |
| *Locura/demente/psicópata* | Craziness/loony/deranged | 13.9 |
| *Olvido/pérdida de memoria* | Forgetfulness/memory loss | 12.4 |
| *Envejecer* | Aging | 9.5 |
| *Tu mente borra* | Erased mind | 9.5 |
| *Confusion/se le va la onda* | Confusion/out of it | 5.6 |
| *No sé* | Don't know | 4.0 |
| *Cerebro se hace pequeño* | Brain gets small | 1.5 |
| *Malvado/perverso* | Evil/wicked | 1.5 |
| *Agresividad* | Aggressive | 0.5 |
| *Depresivo* | Depressive | 0.5 |
| **Words for inherited, k = 145** |  |  |
| *De familia* | Family | 22.8 |
| *En tus genes* | In your genes | 12.4 |
| *Transmisión de características físicas/viene de familia* | Transmission of physical characteristics/comes from family | 11.7 |
| *En la sangre* | In the blood | 4.8 |
| *Nace con eso* | Born with it | 3.4 |
| *No sé* | Do not know | 2.1 |
| *Tierras/dinero/propiedad* | Land/money/property | 2.1 |
| *No se hurta* | Not stolen | 2.1 |
| *No es contagiar* | Not contagious | 1.4 |
| **Words for genes, k = 191** |  |  |
| *Herencia/se transmiten* | Hereditary/are transmitted | 21.5 |
| *La familia/los ancestros* | Family/ancestor | 19.4 |
| *En la sangre* | In the blood | 14.1 |
| *Características/rasgos* | Characteristics/traits | 7.8 |
| *ADN* | DNA | 6.3 |
| *Desde el nacimiento* | Since birth | 3.7 |
| *Células* | Cells | 0.5 |
| **Illnesses worse than AD, k = 132** |  |  |
| *Cáncer* | Cancer | 19.7 |
| *Enfermedades mentales* | Mental illness | 11.4 |
| *Enfermedad de Alzheimer* | Alzheimer's disease | 7.6 |
| *Diabetes* | Diabetes | 6.8 |
| *Enfermedades de los pulmones* | Lung disease | 4.5 |
| *COVID* | COVID | 4.5 |
| *Ataque al corazón* | Heart attack/disease | 4.5 |
| *Síndromes genéticos* | Genetic condition | 3.8 |
| *Derrame cerebral* | Stroke | 0.7 |
| *Vitíligo* | Vitiligo | 0.7 |
| **Causes of AD, k = 179** |  |  |
| *Envejecer* | Aging | 14.0 |
| *Genes* | Genes | 10.6 |
| *La dieta/ nutrición/estilo de vida* | Diet/nutrition/lifestyle | 10.6 |
| *No sé* | Do not know | 6.1 |
| *Deteriorando* | Deterioration/decay | 3.3 |
| *No es una persona activa/ falta de ejercicio* | Physical inactivity | 3.3 |
| *No estimulado mentalmente/interactuó con menos gente* | No mental stimulation/social isolation | 3.3 |
| *Contusión/golpe en la cabeza* | Brain injury | 2.2 |
| *Mucho estrés físico/ nuestro cuerpo ya no es* | Stress on/failure of body | 1.7 |
| Reasons to have AD genetic test, k = 214 |  |  |
| *Si un padre o familiar lo tiene* | If parent or family member has it | 15.9 |
| *Olviden las cosas* | Forgetting | 13.1 |
| *No sé* | Do not know | 6.5 |
| *Prepararse/dejar asuntos en orden/planear nuestro cuidado* | To prepare or plan | 4.2 |
| *Si veo alguien de mi edad que lo tenga* | When someone you know (not a family member) has Alzheimer's | 4.2 |
| *Prevención/medidas preventivas* | Prevention | 3.7 |
| *Nuestra propensión/saber lo que te espera en el futuro* | To determine predisposition | 3.7 |
| *Nuestros hijos* | For children | 2.3 |
| *Curiosidad* | Curiosity | 0.9 |
| Reasons not to have AD genetic test, k = 142 |  |  |
| *No se te olvida/ no tienes ninguna pérdida de memoria/tu mente está bien* | Memory is fine | 13.4 |
| *Miedo saber* | Afraid | 9.9 |
| *No quiero saber/ no quisiera cambiar mi estilo de vida* | Do not want to know/do not care/would not change lifestyle | 6.3 |
| *Ha de ser un proceso caro/ no tiene los recursos* | Cost/expense | 5.6 |
| *Ningún familiar ah ha tenido* | No relatives with disease | 5.6 |
| *Causa angustia/empezar a mortificar* | Causes<br>anguish/shame/embarrassment | 3.5 |
| *No hay alguien que los guíe/ignorancia* | No guidance/ignorance | 2.1 |
| *No sé* | Do not know | 2.1 |
| *No hay tratamiento/no hay nada que hacer* | No treatment/nothing to do | 2.1 |
| **More useful tests than AD genetic test, k = 207** |  |  |
| *Prueba de memoria* | Memory test | 10.1 |
| *Chequeo médico* | Medical evaluation | 6.8 |
| *No sé* | Do not know | 6.3 |
| *Scan del cerebro* | Brain scan | 4.8 |
| *Evaluaciones psicológicas* | Psychological evaluation | 3.9 |
| *Prueba de sangre* | Blood test | 2.9 |
| *Una observación de parte de los familiares* | Others' observations | 1.9 |

### 3.3 Participants know AD as a memory disorder

When asked about “AD,” participants most frequently (33%) commented about “forgetfulness/memory loss” *(olvido/pérdida de memoria)*, nearly 4 times more frequent than the next theme. The concept of AD as a memory disorder appeared to be common knowledge. Additionally, a minority of respondents (6%) listed “craziness/loony/psychopath” *(locura/demente/psicópata)*, consistent with beliefs that AD is a behavioral or psychiatric condition.

### 3.4 “Inherited” and “genes” have cultural salience

Themes from the “inherited” and “genes” prompts reflected familiarity with discrete hereditary units passed from generation to generation. These prompts’ most frequent themes—“family” (*de familia)* (23%) and “hereditary/are transmitted” (*herencia/se transmiten)* (22%)—represented the second and third most frequent themes overall. This suggested these concepts hold broad cultural salience, with minimal variation in discussion. A minority of respondents (2%) associated the non-medical use of “inherited” with bequeathing material items, reflected by the theme “land/money/property” *(tierras/dinero/propiedad)*.

### 3.5 AD genetic testing explains the past/present rather than prepares for the future

When asked about “causes of AD,” the top themes identified were “aging” *(envejecer)* (14%), “genes” *(genes*) (11%), and “diet/nutrition/lifestyle” *(la dieta/ nutrición/estilo de vida)* (11%). Although participants perceived diet/lifestyle and genes as contributing equally—suggesting awareness that AD could occur without family history—they also implied an isolated occurrence of AD did not warrant genetic testing. When prompted for “reasons to have an AD gene test,” top themes were “if parent or family member has it” *(si un padre o familiar lo tiene) (16%)* and “forgetting” *(olviden las cosas) (13%)*. These suggested participants view genetic testing as a tool for understanding past or present health, not forecasting future risk. If understood as solely diagnostic, testing may be perceived as offering limited value for long-term planning. Future-oriented motivations appeared in a minority of responses: “prevention” *(prevención/medidas preventivas)* and “to determine predisposition” *(nuestra propensión/saber lo que te espera en el futuro)* (4% each).

By contrast, the theme “memory is fine” (*No se te olvida/ no tienes ninguna pérdida de memoria/tu mente está bien*) (14%) most frequently characterized “reasons not to have an AD gene test,” followed by “afraid” *(miedo saber)* (10%), highlighting how emotion might also influence decision-making. Other themes, such as “do not want to know/do not care/would not change lifestyle” *(no quiero saber/ no quisiera cambiar mi estilo de vida)* (6%), suggested a test with a direct, tangible impact is more meaningful than one with no actionable information. The theme “no relatives with disease” *(ningún familiar ah ha tenido)* (6%) implied salience of the idea that genetic testing could not be uncoupled from family history, despite the recognition from other themes that AD could occur without affected relatives. While genetic testing for AD seemed pertinent to many, participants also recognized the role of other medical procedures. Themes of “memory test” *(prueba de memoria)* and “medical evaluation” *(chequeo médico)* emerged from the question about “more useful tests than an AD gene test.”

### 3.6 Return of genetic results to individuals with AD invokes sadness and fear

Besides the word/phrase prompts, participants were presented with vignettes in which an individual receives AD genetic results (Table 4). When asked about reactions of the fictional H/L man, participants frequently described emotional responses. Feelings of “sadness/depression” (*estaría triste/se va a poner depresivo* and *triste/deprimido)* were noted for all vignettes about the man with AD: (1) “Rodriguez,” receiving a result confirming autosomal dominant AD (7%); (2) “Mendoza,” receiving a result excluding monogenic AD cause but identifying a hereditary cancer variant (9%); and (3) “Ochoa,” receiving a result excluding monogenic AD but identifying altered medication metabolism (13%). Only the vignette about the man without memory problems (“Enriquez”) did not yield “sad/depressed.” Themes of fear emerged from all vignettes without a high penetrance AD variant: (1) “Enriquez,” “concerned/afraid/worried” *(preocupado/miedo)* (12%); (2) “Mendoza,” “fear/concern” *(miedo/preocupación)* (17%); and (3) “Ochoa,” “worried” (3%). Only the “Rodriguez” vignette did not yield a theme of fear. These data underscore the impact of an identifiable single cause of AD on modulating sadness and fear.

**Table 4.**
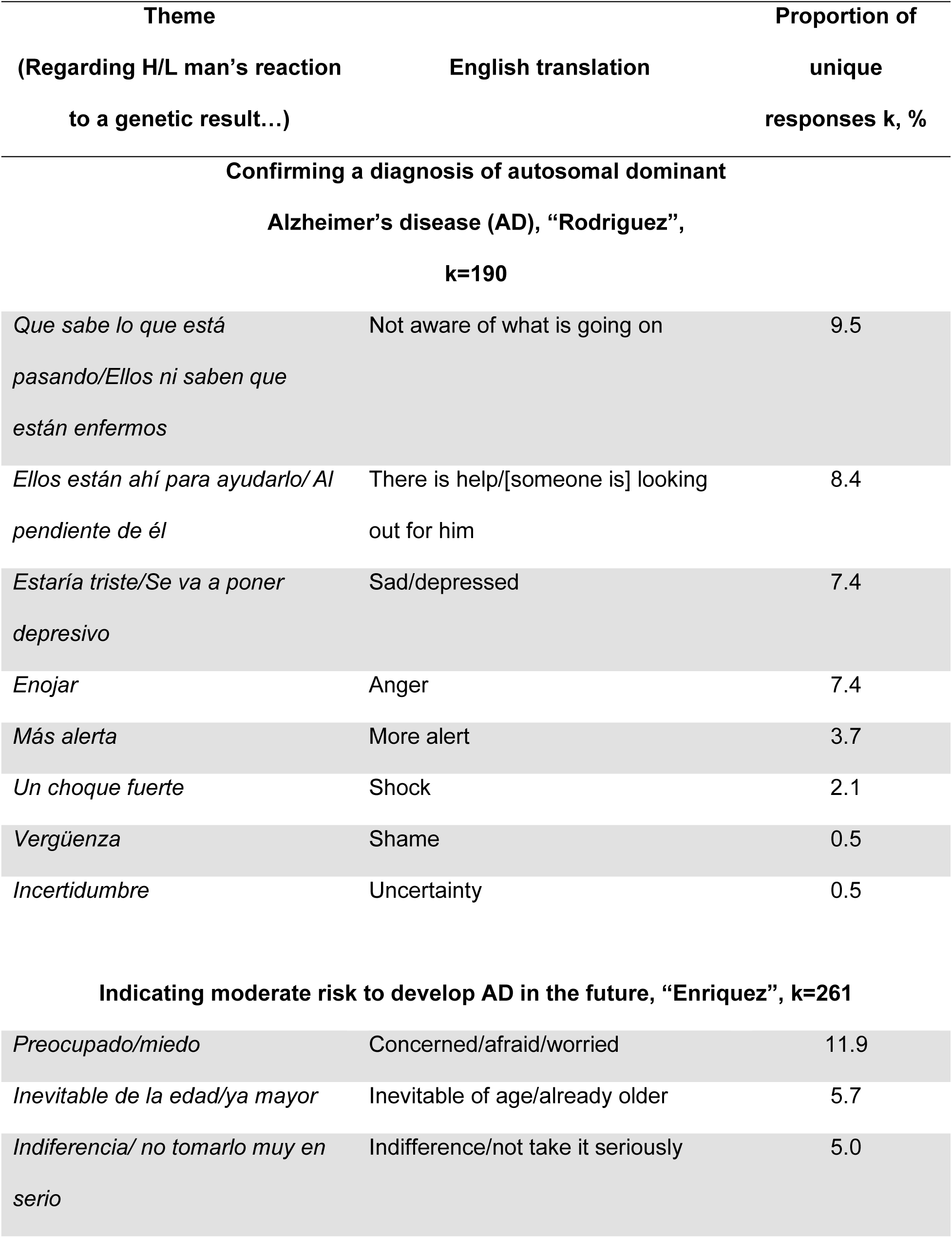
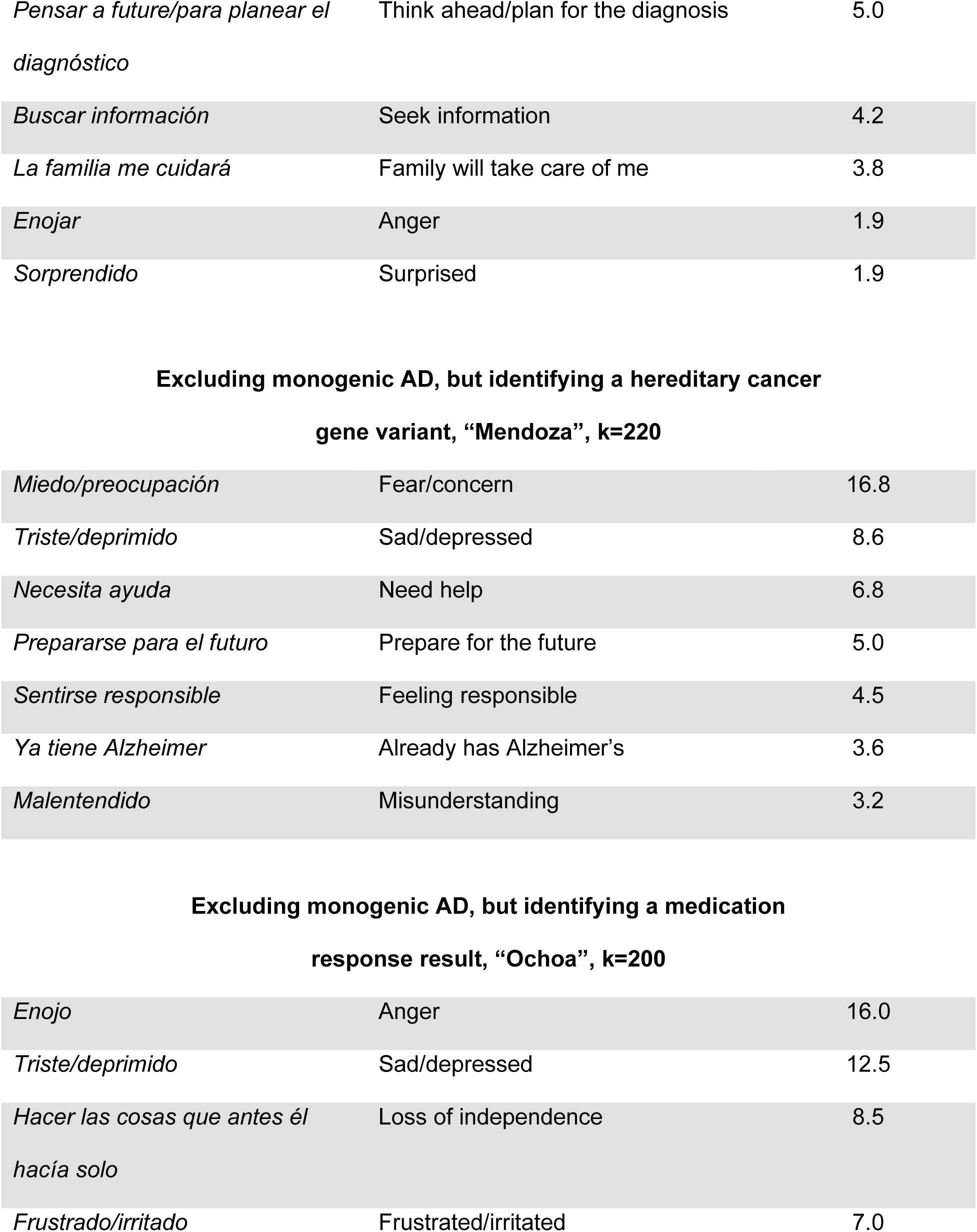
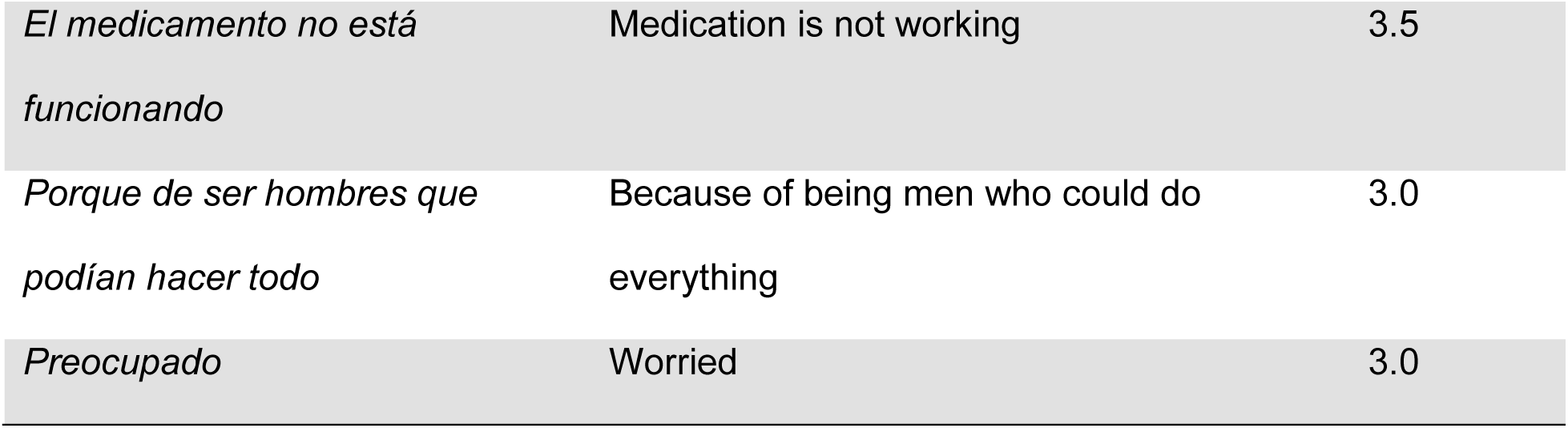
Results of narrative vignette prompts.

| Theme<br>(Regarding H/L man's reaction<br>to a genetic result...) | English translation | Proportion of<br>unique<br>responses k, % |
| --- | --- | --- |
| **Confirming a diagnosis of autosomal dominant**<br>**Alzheimer's disease (AD), "Rodriguez",**<br>**k=190** |  |  |
| *Que sabe lo que está*<br>*pasando/Ellos ni saben que*<br>*están enfermos* | Not aware of what is going on | 9.5 |
| *Ellos están ahí para ayudarlo/ Al*<br>*pendiente de él* | There is help/[someone is] looking<br>out for him | 8.4 |
| *Estaría triste/Se va a poner*<br>*depresivo* | Sad/depressed | 7.4 |
| *Enojar* | Anger | 7.4 |
| *Más alerta* | More alert | 3.7 |
| *Un choque fuerte* | Shock | 2.1 |
| *Vergüenza* | Shame | 0.5 |
| *Incertidumbre* | Uncertainty | 0.5 |
| **Indicating moderate risk to develop AD in the future, "Enriquez", k=261** |  |  |
| *Preocupado/miedo* | Concerned/afraid/worried | 11.9 |
| *Inevitable de la edad/ya mayor* | Inevitable of age/already older | 5.7 |
| *Indiferencia/ no tomarlo muy en*<br>*serio* | Indifference/not take it seriously | 5.0 |
| *Pensar a future/para planear el diagnóstico* | Think ahead/plan for the diagnosis | 5.0 |
| *Buscar información* | Seek information | 4.2 |
| *La familia me cuidará* | Family will take care of me | 3.8 |
| *Enojar* | Anger | 1.9 |
| *Sorprendido* | Surprised | 1.9 |
| **Excluding monogenic AD, but identifying a hereditary cancer gene variant, “Mendoza”, k=220** |  |  |
| *Miedo/preocupación* | Fear/concern | 16.8 |
| *Triste/deprimido* | Sad/depressed | 8.6 |
| *Necesita ayuda* | Need help | 6.8 |
| *Prepararse para el futuro* | Prepare for the future | 5.0 |
| *Sentirse responsable* | Feeling responsible | 4.5 |
| *Ya tiene Alzheimer* | Already has Alzheimer’s | 3.6 |
| *Malentendido* | Misunderstanding | 3.2 |
| **Excluding monogenic AD, but identifying a medication response result, “Ochoa”, k=200** |  |  |
| *Enojo* | Anger | 16.0 |
| *Triste/deprimido* | Sad/depressed | 12.5 |
| *Hacer las cosas que antes él*<br>*hacía solo* | Loss of independence | 8.5 |
| *Frustrado/irritado* | Frustrated/irritated | 7.0 |
| *El medicamento no está funcionando* | Medication is not working | 3.5 |
| *Porque de ser hombres que podían hacer todo* | Because of being men who could do everything | 3.0 |
| *Preocupado* | Worried | 3.0 |

### 3.7 Weakly predictive genetic results engender either a passive or active stance

The ”Enriquez” vignette—describing a man without memory problems learning his (qualitative) AD risk was moderate—elicited “inevitable of age/already older” *(inevitable de la edad/ya mayor)* (6%) and “indifference/not take it seriously” *(indiferencia/ no tomarlo muy en serio)* (5%). These suggest a passive stance toward genetic information: risk is acknowledged but not seen as actionable in the absence of certainty. However, an active stance—interpreting risk as a prompt to plan, prepare, and change behavior—was also salient, as highlighted by “think ahead/plan for the diagnosis” *(pensar a future/para planear el diagnóstico)* (5%), “seek information” *(buscar información)* (4%), and “family will take care of me” *(la familia me cuidará)* (4%).

### 3.8 Return of non-AD risk results generated unanticipated responses

The “Mendoza” vignette—describing a man with AD learning that he lacked a high penetrance AD gene variant but was predisposed for cancer—elicited “misunderstanding” *(malentendido)* (3%). No other vignette generated similar themes, suggesting a minority of participants did not anticipate genetic results relevant to other health conditions.

The “Ochoa” vignette—describing a man with AD receiving a medication response result—elicited the theme “because of being men who could do everything” (*porque de ser hombres que podían hacer todo)* (3%). The scenario described the man’s irritability and suboptimal anti-depressant treatment because he was an ultra-rapid metabolizer, as well as his distress at losing independence. Description of the man in distress prompted respondents to identify prideful masculinity as salient, though whether this stemmed from return of genetic results or from functional decline could not be determined.

## 4. DISCUSSION

This study reveals the words that Spanish preferring Mexican/Mexican American adults in Houston use to talk about AD and genetics as well as perceptions about genetic testing in the context of AD. Participants were generally familiar with both “AD” and “dementia,” but the latter term was more likely to be associated with mental illness. Although few studies have focused on perceptions among H/L individuals, this result is consistent with other published work suggesting more limited awareness of the behavioral and neuropsychiatric manifestations of AD.^20,31,32^ Our results also add to prior studies highlighting awareness among H/L adults that AD is a multifactorial disorder with important contribution from genes along with aging and other lifestyle choices/behaviors that may impact brain health.^20,32,33^ We documented a high level of overall interest in AD genetic testing, and greater proximity to the disease influenced the desire to have such testing (e.g., if either the participant or their friends/family were displaying symptoms). While most respondents were familiar with the idea that biological traits can be transmitted across generations, the word “inherited” was associated more with “property” or “something bequeathed,” suggesting the need for clearer explanation or alternative wording in AD education models. Nevertheless, results from our freelisting interviews are consistent with other studies indicating that many H/L adults are already aware that genetic testing can reveal disease risk, including for themselves and their families, and may also inform disease treatment.^34,35^

On average, participants reported they would be very likely to participate in clinical research involving AD genetic testing, even though some were uncertain about its personal relevance or utility. This enthusiasm may reflect perceptions of a technological imperative surrounding genetic testing.^36^ Prior research suggests that the availability of medical technology for people with or at risk for hereditary neurodegenerative disease creates pressure to use it. Declining such technology may provoke feelings of guilt, irresponsibility, and self-blame.^37,38^ Exploring whether advances in AD genetic knowledge creates a feeling of responsibility to pursue genetic testing—despite poor understanding of the test’s purpose—may inform strategies for H/L education and outreach. For example, explanatory materials might include not only information about what AD genetic testing is, but also concrete examples of what people might do with the information, as well as limitations.

Importantly, our results also identify several potential barriers to universal acceptance of AD genetic testing by Spanish-preferring H/L adults. Unexpectedly, respondents cited intact memory as a reason not to have AD genetic testing, consistent with a belief that testing may be useful solely for diagnostic, rather than predictive purposes. The idea that cognitively normal individuals might seek genetic testing to anticipate future AD risk—using this information to plan for future care or finances—did not emerge as a salient theme among respondents. Instead, participants tended to view genetic testing as appropriate only when cognitive symptoms were already present, to inform diagnosis or treatment decisions. This contrast reflects a dispositional orientation toward present health, emphasizing action only in response to current symptoms rather than to prepare for potential future risk. Consequently, educational materials might emphasize how genetic testing of cognitively normal individuals can estimate AD risk, potentially motivating lifestyle changes (e.g., diet and exercise) that promote brain health and possibly prevent future decline—altogether reflecting a dispositional orientation toward future health. In addition, our study revealed that many individuals lack understanding of the potential for secondary findings during genetic testing, making information about both primary and secondary findings important to include in educational materials. Current guidelines recommend returning secondary, medically-actionable findings from clinical genetic testing.^39^ However, few studies have examined pertinent beliefs among patients and caregivers in the context of clinical care and counseling for AD genetic testing. In addition to its potential impact on future clinical care, the disclosure of individualized actionable findings is recognized as a key motivator for participation in genomic research—contributing to both recruitment and retention,^40–43^ including among individuals from underrepresented groups such as H/L adults.

Our results, combined with findings from similar studies, can powerfully inform initiatives to improve communication with Spanish-preferring H/L adults about AD and genetic testing in the future, ensuring better-informed decisions for patients and their family members. Community-tailored communication strategies should also be developed in partnership with H/L community members and advisors. In addition to guiding the development of print and digital materials, these findings highlight the need to enhance training of physicians, genetic counselors, and other members of clinical care and research teams to deliver culturally relevant information and to engage the H/L community in meaningful and effective ways. Although genetic counseling is well established in genetics clinics and neurology practices at many academic centers caring for patients with familial AD and other neurodegenerative diseases with strong genetic contributions,^44,45^ many clinicians who care for older adults at risk for AD may be ill-equipped to discuss genetics results and their implications, particularly with Spanish-preferring H/L individuals. Our participants regarded *familismo*, a H/L cultural value characterized by a high degree of social interaction, obligation, and support among nuclear and extended family members,^46,47^ as highly important.

Accordingly, AD genetic testing outreach may be strengthened by emphasizing that test results can inform and potentially benefit relatives, aligning with familial priorities and support systems. Prior studies have documented that deep spiritual and religious beliefs can be integrated into the daily lives of individuals from many H/L cultures^48–50^ potentially influencing perceptions of health and aging. Interestingly, we did not identify themes of spirituality in responses from this study, possibly because participants were primed to think about biological factors, based on the order of presentation and nature of the interview prompts and vignettes.

Another potential limitation of our study is that our findings could be restricted to the experiences of our participants, all of whom identified as Mexican or Mexican American. Follow-up confirmatory studies in other H/L heritage groups may be important to establish generalizability. In addition, freelisting is one qualitative data collection strategy. It is possible that other approaches would have yielded different themes.

Overall, our findings from this study offer a foundation for developing culturally informed communication strategies, shaped by contributions of both H/L community members and researchers. Effective AD education and outreach, incorporating content that community members identify as salient about genetic testing and its varied results, can help ensure all individuals have an opportunity to engage in and benefit from ongoing advances in AD research.

## Data Availability

All data produced in the present study are available upon reasonable request to the authors.

## Acknowledgements

We would like to thank the participants in this study, as well as the Consulate General of Mexico in Houston, the Literacy Initiative for Today of University of St. Thomas, BakerRipley, and the Alzheimer’s Association Houston and Southeast Texas Chapter. We also thank Cynthia Montiel-Castillo for her assistance with data curation.

## Consent statement

The authors obtained study approval from the Institutional Review Board at Baylor College of Medicine (H-49633). All participants provided informed consent.

## Supplemental Material

**Supplemental Table 1.**
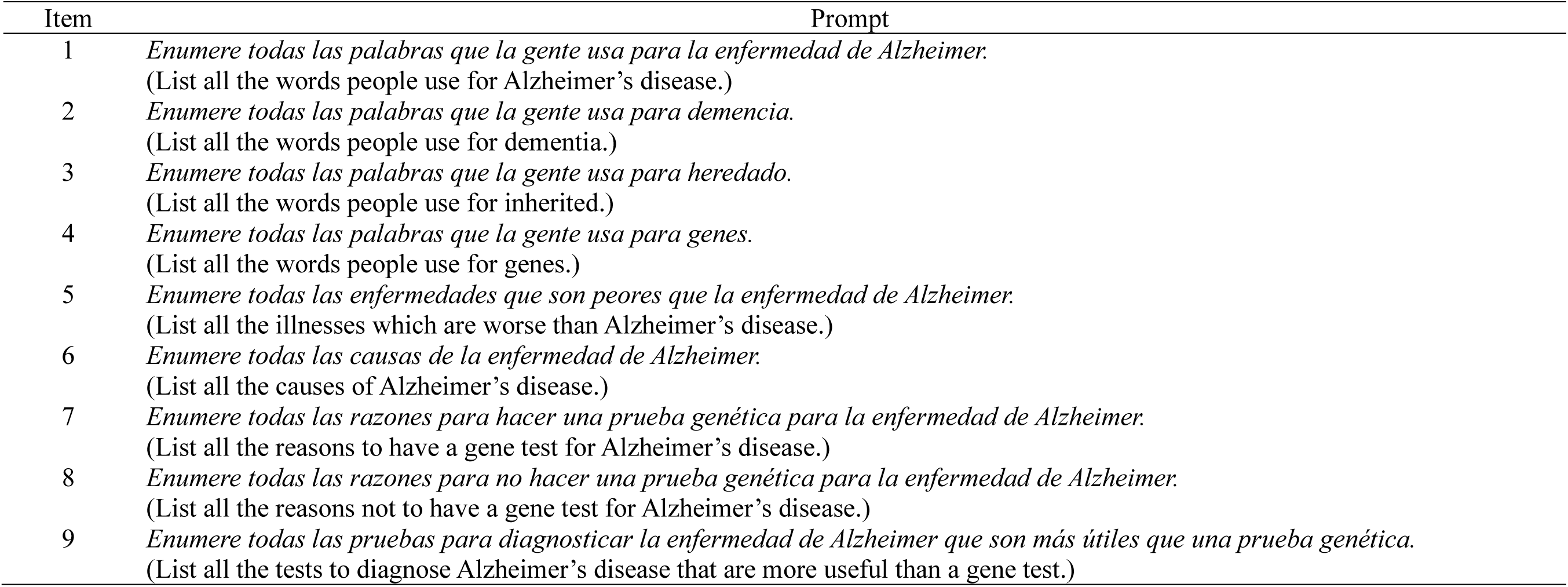
Word cluster prompts comprising the freelisting interview with each participant.

**Supplemental Table 2.**
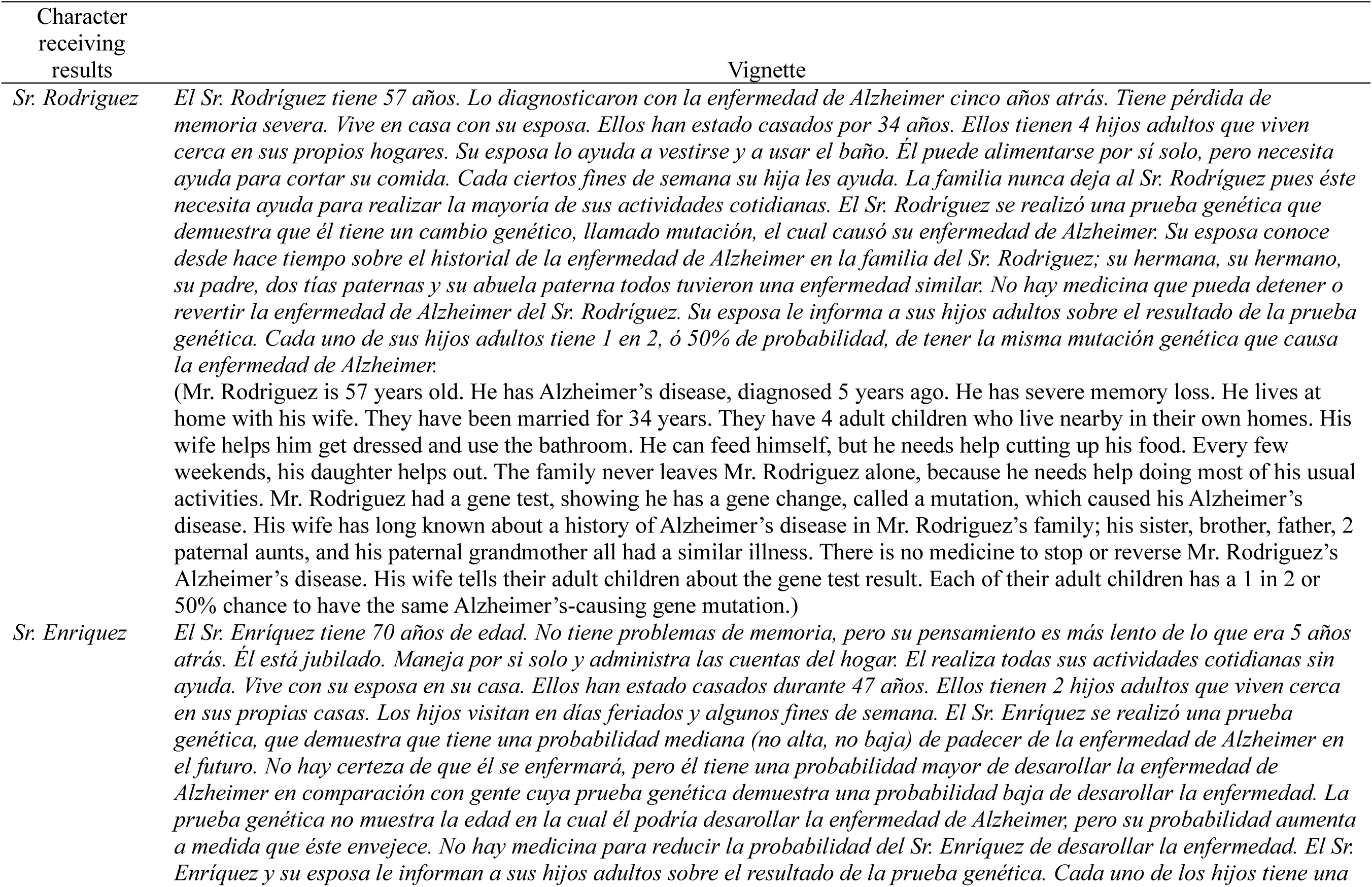
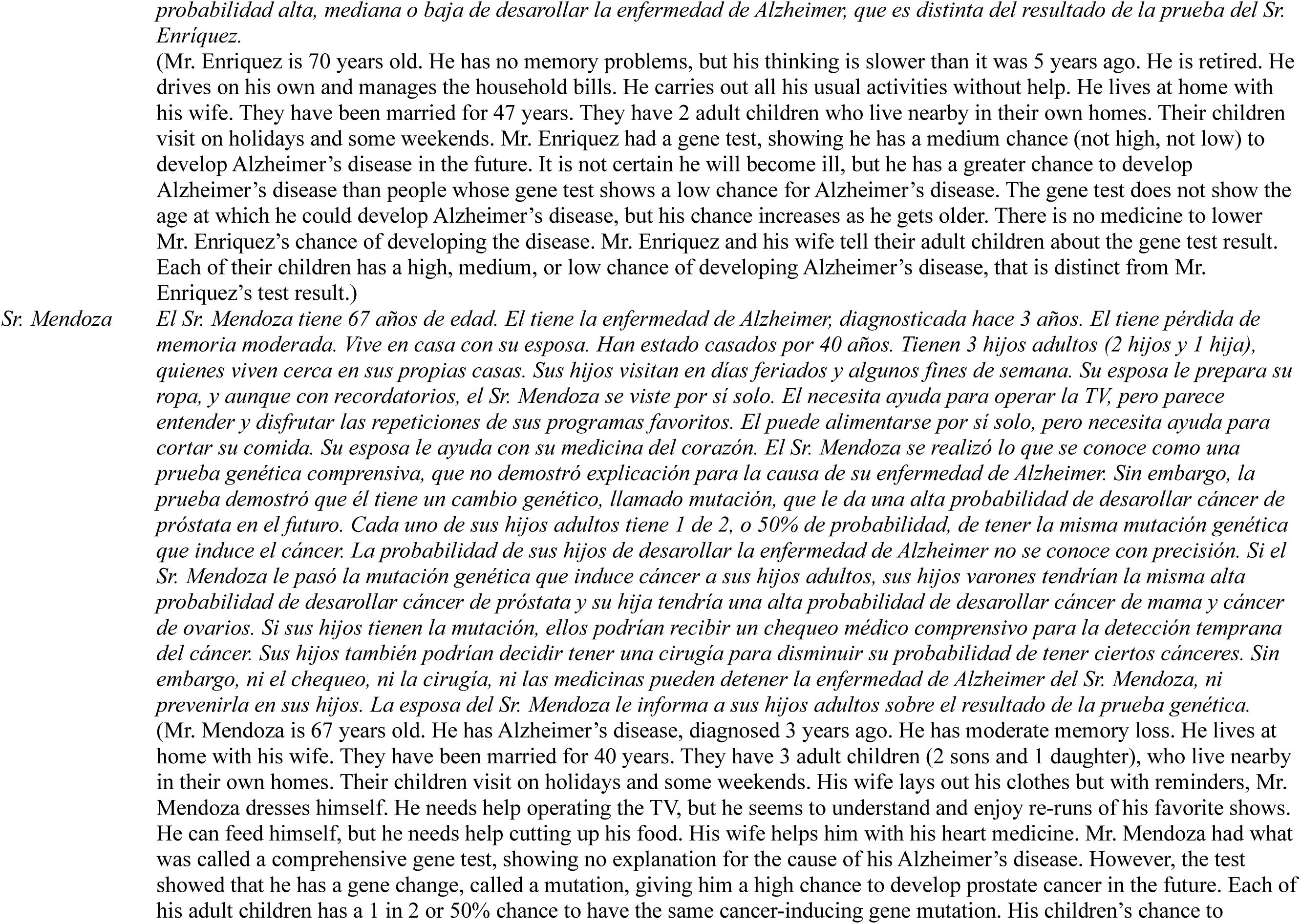
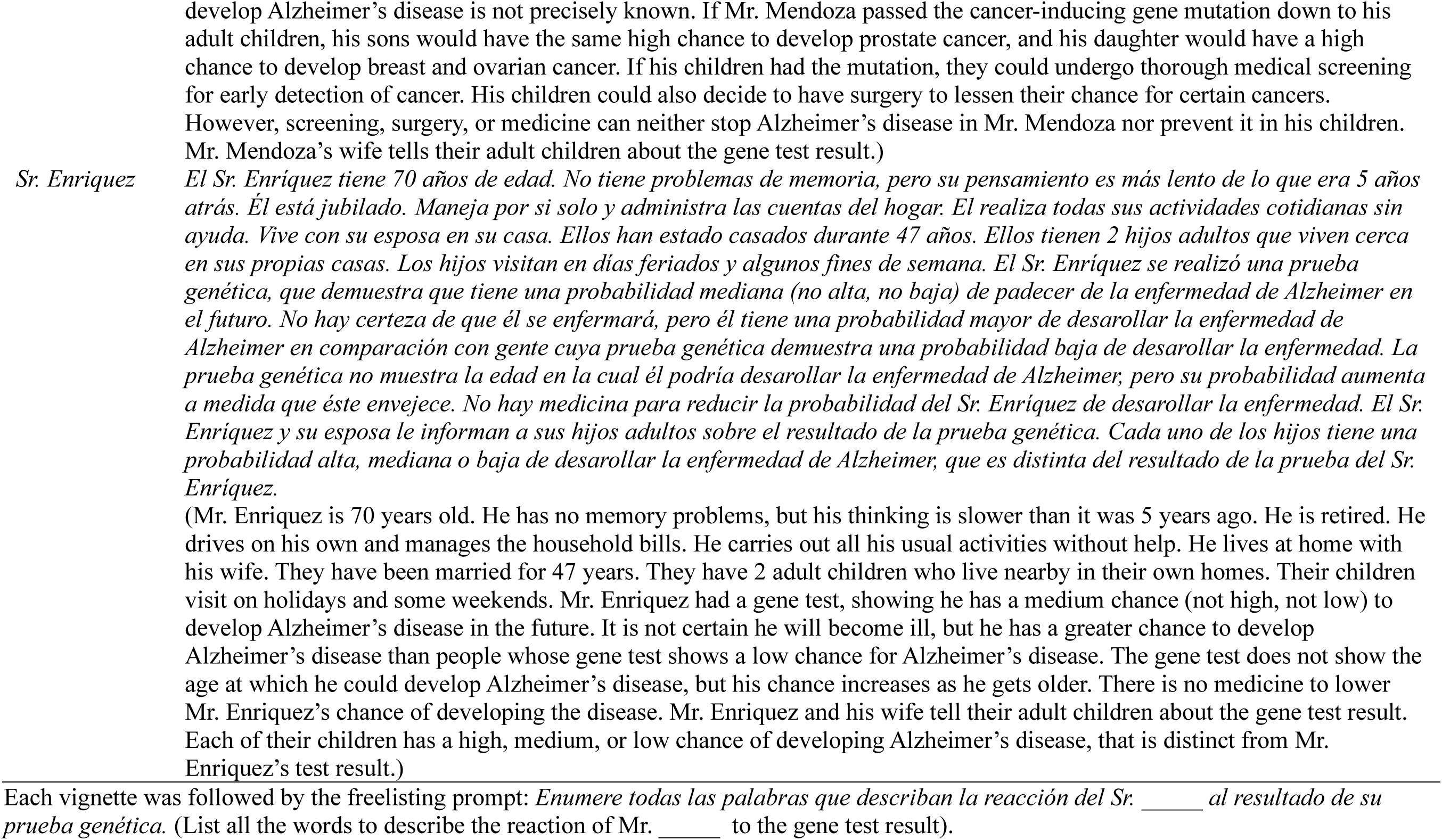
Vignettes comprising the freelisting interview with each participant.

## References

1. Gurland BJ, Wilder DE, Lantigua R, et al. Rates of dementia in three ethnoracial groups. International Journal of Geriatric Psychiatry 1999; 14: 481–493.

2. Haan MN, Mungas DM, Gonzalez HM, et al. Prevalence of dementia in older Latinos: The influence of type 2 diabetes mellitus, stroke and genetic factors. Journal of the American Geriatrics Society 2003; 51: 169–177.

3. Samper-Ternent R, Kuo YF, Ray LA, et al. Prevalence of health conditions and predictors of mortality in oldest old Mexican Americans and non-Hispanic whites. Journal of the American Medical Directors Association 2012; 13: 254–259.

4. Bellenguez C, Küçükali F, Jansen IE, et al. New insights into the genetic etiology of Alzheimer’s disease and related dementias. Nat Genet 2022; 54: 412–436.

5. Jansen IE, Savage JE, Watanabe K, et al. Genome-wide meta-analysis identifies new loci and functional pathways influencing Alzheimer’s disease risk. Nature Genetics 2019; 51: 404–413.

6. Kunkle BW, Grenier-Boley B, Sims R, et al. Genetic meta-analysis of diagnosed Alzheimer’s disease identifies new risk loci and implicates Aβ, tau, immunity and lipid processing. Nat Genet 2019; 51: 414–430.

7. Adkins-Jackson PB, George KM, Besser LM, et al. The structural and social determinants of Alzheimer’s disease related dementias. Alzheimer’s & Dementia 2023; 19: 3171–3185.

8. Coogan P, Schon K, Li S, et al. Experiences of racism and subjective cognitive function in African American women. Alzheimer’s & Dementia: Diagnosis, Assessment & Disease Monitoring 2020; 12: e12067.

9. González HM, Tarraf W, Jian X, et al. Apolipoprotein E genotypes among diverse middle-aged and older Latinos: Study of Latinos-Investigation of Neurocognitive Aging results (HCHS/SOL). Sci Rep 2018; 8: 17578.

10. Dickerson BC, Atri A, Clevenger C, et al. The Alzheimer’s Association clinical practice guideline for the Diagnostic Evaluation, Testing, Counseling, and Disclosure of Suspected Alzheimer’s Disease and Related Disorders (DETeCD-ADRD): Executive summary of recommendations for specialty care. Alzheimer’s & Dementia; n/a. DOI: 10.1002/alz.14337.

11. Cummings J, Apostolova L, Rabinovici GD, et al. Lecanemab: Appropriate use recommendations. The Journal of Prevention of Alzheimer’s Disease 2023; 10: 362–377.

12. Rabinovici GD, Selkoe DJ, Schindler SE, et al. Donanemab: Appropriate use recommendations. The Journal of Prevention of Alzheimer’s Disease 2025; 12: 100150.

13. Mayeda ER, Glymour MM, Quesenberry CP, et al. Inequalities in dementia incidence between six racial and ethnic groups over 14 years. Alzheimer’s & Dementia 2016; 12: 216–224.

14. Wilkins CH, Schindler SE, Morris JC. Addressing health disparities among minority populations: Why clinical trial recruitment is not enough. JAMA Neurology 2020; 77: 1063–1064.

15. Arce Rentería M, Mobley TM, Evangelista ND, et al. Representativeness of samples enrolled in Alzheimer’s disease research centers. Alzheimer’s & Dementia: Diagnosis, Assessment & Disease Monitoring 2023; 15: e12450.

16. Migration Policy Institute. State Language Data - US. migrationpolicy.org, https://www.migrationpolicy.org/data/state-profiles/state/language/US (accessed 11 October 2023).

17. Dron HA, Bucio D, Young JL, et al. Latinx attitudes, barriers, and experiences with genetic counseling and testing: A systematic review. J Genet Couns 2023; 32: 166–181.

18. Hamilton J, Shuk E, Arniella G, et al. Genetic testing awareness and attitudes among Latinos: Exploring shared perceptions and gender-based differences. Public health genomics 2016; 19: 34– 46.

19. Greater Houston Community Foundation. Population, diversity, and immigration in Houston, https://www.understandinghouston.org/topic/community-context/population-and-diversity#overview (accessed 31 July 2025).

20. Karlawish J, Barg FK, Augsburger D, et al. What Latino Puerto Ricans and non-Latinos say when they talk about Alzheimer’s disease. Alzheimer’s & Dementia 2011; 7: 161–170.

21. Irby MB, Moore KR, Mann-Jackson L, et al. Community-engaged research: Common themes and needs identified by investigators and research teams at an emerging academic learning health system. Int J Environ Res Public Health 2021; 18: 3893.

22. Rhodes SD, Tanner AE, Mann-Jackson L, et al. Promoting community and population health in public health and medicine: A stepwise guide to initiating and conducting community-engaged research. Journal of health disparities research and practice 2018; 11: 16.

23. Wallerstein N, Duran B. Community-based participatory research contributions to intervention research: The intersection of science and practice to improve health equity. Am J Public Health 2010; 100: S40–S46.

24. Marín G, Gamba RJ. A new measurement of acculturation for Hispanics: The bidimensional acculturation scale for Hispanics (BAS). Hisp J Behav Sci 1996; 18: 297–316.

25. Villarreal R, Blozis SA, Widaman KF. Factorial invariance of a pan-Hispanic familism scale. Hisp J Behav Sci 2005; 27: 409–425.

26. Chew LD, Griffin JM, Partin MR, et al. Validation of screening questions for limited health literacy in a large VA outpatient population. J Gen Intern Med 2008; 23: 561–566.

27. Sarkar U, Schillinger D, López A, et al. Validation of self-reported health literacy questions among diverse English and Spanish-speaking populations. J Gen Intern Med 2011; 26: 265–271.

28. Quinlan, M.B. The Freelisting Method. In: Handbook of Research Methods in Health Social Sciences. Singapore: Springer International Publishing, pp. 1–16.

29. Brewer DD. Supplementary interviewing techniques to maximize output in free listing tasks. Field Methods 2002; 14: 108–118.

30. Weller S, Romney A. Systematic Data Collection. SAGE Publications, Inc. Epub ahead of print 1988. DOI: 10.4135/9781412986069.

31. Cabrera LY, Kelly P, Vega IE. Knowledge and attitudes of two Latino groups about Alzheimer disease: A qualitative study. Journal of Cross-Cultural Gerontology 2021; 36: 265–284.

32. Light SW, Tomasino F, Wescott A, et al. Perceptions, beliefs, attitudes, and knowledge of US Latino adults pertaining to dementia and brain health: a systematic review. Aging & Mental Health 2023; 0: 1–12.

33. Ayalon L. Re-examining ethnic differences in concerns, knowledge, and beliefs about Alzheimer’s disease: Results from a national sample. International Journal of Geriatric Psychiatry 2013; 28: 1288–1295.

34. Sorlie PD, Avilés-Santa LM, Wassertheil-Smoller S, et al. Design and implementation of the Hispanic Community Health Study/Study of Latinos. Ann Epidemiol 2010; 20: 629–641.

35. Christensen KD, Zhang M, Galbraith LN, et al. Awareness and utilization of genetic testing among Hispanic and Latino adults living in the US: The Hispanic Community Health Study/Study of Latinos. HGG Adv 2022; 4: 100160.

36. Hofmann B. Is there a technological imperative in health care? International Journal of Technology Assessment in Health Care 2002; 18: 675–89.

37. Adam S, Wiggins S, Whyte P, et al. Five year study of prenatal testing for Huntington’s disease: demand, attitudes, and psychological assessment. J Med Genet 1993; 30: 549–556.

38. Decruyenaere M, Evers-Kiebooms G, Boogaerts A, et al. The complexity of reproductive decision-making in asymptomatic carriers of the Huntington mutation. Eur J Hum Genet 2007; 15: 453–462.

39. Lee K, Abul-Husn NS, Amendola LM, et al. ACMG SF v3.3 list for reporting of secondary findings in clinical exome and genome sequencing: A policy statement of the American College of Medical Genetics and Genomics (ACMG). Genetics in Medicine; 27. Epub ahead of print 1 August 2025. DOI: 10.1016/j.gim.2025.101454.

40. Goodman D, Bowen D, Wenzel L, et al. The research participant perspective related to the conduct of genomic cohort studies: A systematic review of the quantitative literature. Transl Behav Med 2018; 8: 119–129.

41. Gollust SE, Gordon ES, Zayac C, et al. Motivations and perceptions of early adopters of personalized genomics: Perspectives from research participants. Public Health Genomics 2011; 15: 22–30.

42. Kauffman TL, Irving SA, Leo MC, et al. The NextGen Study: Patient motivation for participation in genome sequencing for carrier status. Mol Genet Genomic Med 2017; 5: 508–515.

43. Lupo PJ, Robinson JO, Diamond PM, et al. Patients’ perceived utility of whole-genome sequencing for their healthcare: Findings from the MedSeq project. Per Med 2016; 13: 13–20.

44. Roberts JS, Patterson AK, Uhlmann WR. Genetic testing for neurodegenerative diseases: Ethical and health communication challenges. Neurobiology of Disease 2020; 141: 104871.

45. Dratch L, Azage M, Baldwin A, et al. Genetic testing in adults with neurologic disorders: Indications, approach, and clinical impacts. J Neurol 2024; 271: 733–747.

46. Marín G, Marín BV. Research with Hispanic populations. Newbury Park, Calif.; London: SAGE Publications, Inc., https://methods.sagepub.com/book/research-with-hispanic-populations (1991).

47. Sabogal F, Marín G, Otero-Sabogal R, et al. Hispanic familism and acculturation: What changes and what doesn’t? Hisp J Behav Sci 1987; 9: 397–412.

48. Campesino M, Schwartz GE. Spirituality among Latinas/os: Implications of culture in conceptualization and measurement. ANS Adv Nurs Sci 2006; 29: 69–81.

49. Weiner-Light S, Rankin KP, Lanata S, et al. The role of spirituality in conceptualizations of health maintenance and healthy aging among Latin American immigrants. The American Journal of Geriatric Psychiatry 2021; 29: 1079–1088.

50. Sharkey JR, Sharf BF, St. John JA. “Una persona derechita (staying right in the mind)”: Perceptions of Spanish-speaking Mexican American older adults in South Texas colonias. The Gerontologist 2009; 49: S79–S85.

