## Supplemental Materials for "The words community dwelling, Spanish preferring Hispanic/Latino adults use to talk about Alzheimer’s disease and genetic testing: Implications for education and outreach"

**Supplemental Material**

**Supplemental Table 1.** Word cluster prompts comprising the freelisting interview with each participant.

| Item | Prompt |
|---|---|
| 1 | *Enumere todas las palabras que la gente usa para la enfermedad de Alzheimer.*<br>(List all the words people use for Alzheimer's disease.) |
| 2 | *Enumere todas las palabras que la gente usa para demencia.*<br>(List all the words people use for dementia.) |
| 3 | *Enumere todas las palabras que la gente usa para heredado.*<br>(List all the words people use for inherited.) |
| 4 | *Enumere todas las palabras que la gente usa para genes.*<br>(List all the words people use for genes.) |
| 5 | *Enumere todas las enfermedades que son peores que la enfermedad de Alzheimer.*<br>(List all the illnesses which are worse than Alzheimer's disease.) |
| 6 | *Enumere todas las causas de la enfermedad de Alzheimer.*<br>(List all the causes of Alzheimer's disease.) |
| 7 | *Enumere todas las razones para hacer una prueba genética para la enfermedad de Alzheimer.*<br>(List all the reasons to have a gene test for Alzheimer's disease.) |
| 8 | *Enumere todas las razones para no hacer una prueba genética para la enfermedad de Alzheimer.*<br>(List all the reasons not to have a gene test for Alzheimer's disease.) |
| 9 | *Enumere todas las pruebas para diagnosticar la enfermedad de Alzheimer que son más útiles que una prueba genética.*<br>(List all the tests to diagnose Alzheimer's disease that are more useful than a gene test.) |

**Supplemental Table 2.** Vignettes comprising the freelisting interview with each participant.

| Character receiving results | Vignette |
|---|---|
| *Sr. Rodriguez* | *El Sr. Rodríguez tiene 57 años. Lo diagnosticaron con la enfermedad de Alzheimer cinco años atrás. Tiene pérdida de memoria severa. Vive en casa con su esposa. Ellos han estado casados por 34 años. Ellos tienen 4 hijos adultos que viven cerca en sus propios hogares. Su esposa lo ayuda a vestirse y a usar el baño. Él puede alimentarse por sí solo, pero necesita ayuda para cortar su comida. Cada ciertos fines de semana su hija les ayuda. La familia nunca deja al Sr. Rodríguez pues éste necesita ayuda para realizar la mayoría de sus actividades cotidianas. El Sr. Rodríguez se realizó una prueba genética que demuestra que él tiene un cambio genético, llamado mutación, el cual causó su enfermedad de Alzheimer. Su esposa conoce desde hace tiempo sobre el historial de la enfermedad de Alzheimer en la familia del Sr. Rodriguez; su hermana, su hermano, su padre, dos tías paternas y su abuela paterna todos tuvieron una enfermedad similar. No hay medicina que pueda detener o revertir la enfermedad de Alzheimer del Sr. Rodríguez. Su esposa le informa a sus hijos adultos sobre el resultado de la prueba genética. Cada uno de sus hijos adultos tiene 1 en 2, ó 50% de probabilidad, de tener la misma mutación genética que causa la enfermedad de Alzheimer.*<br>(Mr. Rodriguez is 57 years old. He has Alzheimer’s disease, diagnosed 5 years ago. He has severe memory loss. He lives at home with his wife. They have been married for 34 years. They have 4 adult children who live nearby in their own homes. His wife helps him get dressed and use the bathroom. He can feed himself, but he needs help cutting up his food. Every few weekends, his daughter helps out. The family never leaves Mr. Rodriguez alone, because he needs help doing most of his usual activities. Mr. Rodriguez had a gene test, showing he has a gene change, called a mutation, which caused his Alzheimer’s disease. His wife has long known about a history of Alzheimer’s disease in Mr. Rodriguez’s family; his sister, brother, father, 2 paternal aunts, and his paternal grandmother all had a similar illness. There is no medicine to stop or reverse Mr. Rodriguez’s Alzheimer’s disease. His wife tells their adult children about the gene test result. Each of their adult children has a 1 in 2 or 50% chance to have the same Alzheimer’s-causing gene mutation.) |
| *Sr. Enriquez* | *El Sr. Enríquez tiene 70 años de edad. No tiene problemas de memoria, pero su pensamiento es más lento de lo que era 5 años atrás. Él está jubilado. Maneja por si solo y administra las cuentas del hogar. El realiza todas sus actividades cotidianas sin ayuda. Vive con su esposa en su casa. Ellos han estado casados durante 47 años. Ellos tienen 2 hijos adultos que viven cerca en sus propias casas. Los hijos visitan en días feriados y algunos fines de semana. El Sr. Enríquez se realizó una prueba genética, que demuestra que tiene una probabilidad mediana (no alta, no baja) de padecer de la enfermedad de Alzheimer en el futuro. No hay certeza de que él se enfermará, pero él tiene una probabilidad mayor de desarollar la enfermedad de Alzheimer en comparación con gente cuya prueba genética demuestra una probabilidad baja de desarollar la enfermedad. La prueba genética no muestra la edad en la cual él podría desarollar la enfermedad de Alzheimer, pero su probabilidad aumenta a medida que éste envejece. No hay medicina para reducir la probabilidad del Sr. Enríquez de desarollar la enfermedad. El Sr. Enríquez y su esposa le informan a sus hijos adultos sobre el resultado de la prueba genética. Cada uno de los hijos tiene una* |

*probabilidad alta, mediana o baja de desarollar la enfermedad de Alzheimer, que es distinta del resultado de la prueba del Sr. Enríquez.*
(Mr. Enriquez is 70 years old. He has no memory problems, but his thinking is slower than it was 5 years ago. He is retired. He drives on his own and manages the household bills. He carries out all his usual activities without help. He lives at home with his wife. They have been married for 47 years. They have 2 adult children who live nearby in their own homes. Their children visit on holidays and some weekends. Mr. Enriquez had a gene test, showing he has a medium chance (not high, not low) to develop Alzheimer's disease in the future. It is not certain he will become ill, but he has a greater chance to develop Alzheimer's disease than people whose gene test shows a low chance for Alzheimer's disease. The gene test does not show the age at which he could develop Alzheimer's disease, but his chance increases as he gets older. There is no medicine to lower Mr. Enriquez's chance of developing the disease. Mr. Enriquez and his wife tell their adult children about the gene test result. Each of their children has a high, medium, or low chance of developing Alzheimer's disease, that is distinct from Mr. Enriquez's test result.)

*Sr. Mendoza*

*El Sr. Mendoza tiene 67 años de edad. El tiene la enfermedad de Alzheimer, diagnosticada hace 3 años. El tiene pérdida de memoria moderada. Vive en casa con su esposa. Han estado casados por 40 años. Tienen 3 hijos adultos (2 hijos y 1 hija), quienes viven cerca en sus propias casas. Sus hijos visitan en días feriados y algunos fines de semana. Su esposa le prepara su ropa, y aunque con recordatorios, el Sr. Mendoza se viste por sí solo. El necesita ayuda para operar la TV, pero parece entender y disfrutar las repeticiones de sus programas favoritos. El puede alimentarse por sí solo, pero necesita ayuda para cortar su comida. Su esposa le ayuda con su medicina del corazón. El Sr. Mendoza se realizó lo que se conoce como una prueba genética comprensiva, que no demostró explicación para la causa de su enfermedad de Alzheimer. Sin embargo, la prueba demostró que él tiene un cambio genético, llamado mutación, que le da una alta probabilidad de desarollar cáncer de próstata en el futuro. Cada uno de sus hijos adultos tiene 1 de 2, o 50% de probabilidad, de tener la misma mutación genética que induce el cáncer. La probabilidad de sus hijos de desarollar la enfermedad de Alzheimer no se conoce con precisión. Si el Sr. Mendoza le pasó la mutación genética que induce cáncer a sus hijos adultos, sus hijos varones tendrían la misma alta probabilidad de desarollar cáncer de próstata y su hija tendría una alta probabilidad de desarollar cáncer de mama y cáncer de ovarios. Si sus hijos tienen la mutación, ellos podrían recibir un chequeo médico comprensivo para la detección temprana del cáncer. Sus hijos también podrían decidir tener una cirugía para disminuir su probabilidad de tener ciertos cánceres. Sin embargo, ni el chequeo, ni la cirugía, ni las medicinas pueden detener la enfermedad de Alzheimer del Sr. Mendoza, ni prevenirla en sus hijos. La esposa del Sr. Mendoza le informa a sus hijos adultos sobre el resultado de la prueba genética.*
(Mr. Mendoza is 67 years old. He has Alzheimer's disease, diagnosed 3 years ago. He has moderate memory loss. He lives at home with his wife. They have been married for 40 years. They have 3 adult children (2 sons and 1 daughter), who live nearby in their own homes. Their children visit on holidays and some weekends. His wife lays out his clothes but with reminders, Mr. Mendoza dresses himself. He needs help operating the TV, but he seems to understand and enjoy re-runs of his favorite shows. He can feed himself, but he needs help cutting up his food. His wife helps him with his heart medicine. Mr. Mendoza had what was called a comprehensive gene test, showing no explanation for the cause of his Alzheimer's disease. However, the test showed that he has a gene change, called a mutation, giving him a high chance to develop prostate cancer in the future. Each of his adult children has a 1 in 2 or 50% chance to have the same cancer-inducing gene mutation. His children's chance to

| | |
|---|---|
| | develop Alzheimer's disease is not precisely known. If Mr. Mendoza passed the cancer-inducing gene mutation down to his adult children, his sons would have the same high chance to develop prostate cancer, and his daughter would have a high chance to develop breast and ovarian cancer. If his children had the mutation, they could undergo thorough medical screening for early detection of cancer. His children could also decide to have surgery to lessen their chance for certain cancers. However, screening, surgery, or medicine can neither stop Alzheimer's disease in Mr. Mendoza nor prevent it in his children. Mr. Mendoza's wife tells their adult children about the gene test result.) |
| *Sr. Enriquez* | *El Sr. Enríquez tiene 70 años de edad. No tiene problemas de memoria, pero su pensamiento es más lento de lo que era 5 años atrás. Él está jubilado. Maneja por si solo y administra las cuentas del hogar. El realiza todas sus actividades cotidianas sin ayuda. Vive con su esposa en su casa. Ellos han estado casados durante 47 años. Ellos tienen 2 hijos adultos que viven cerca en sus propias casas. Los hijos visitan en días feriados y algunos fines de semana. El Sr. Enríquez se realizó una prueba genética, que demuestra que tiene una probabilidad mediana (no alta, no baja) de padecer de la enfermedad de Alzheimer en el futuro. No hay certeza de que él se enfermará, pero él tiene una probabilidad mayor de desarollar la enfermedad de Alzheimer en comparación con gente cuya prueba genética demuestra una probabilidad baja de desarollar la enfermedad. La prueba genética no muestra la edad en la cual él podría desarollar la enfermedad de Alzheimer, pero su probabilidad aumenta a medida que éste envejece. No hay medicina para reducir la probabilidad del Sr. Enríquez de desarollar la enfermedad. El Sr. Enríquez y su esposa le informan a sus hijos adultos sobre el resultado de la prueba genética. Cada uno de los hijos tiene una probabilidad alta, mediana o baja de desarollar la enfermedad de Alzheimer, que es distinta del resultado de la prueba del Sr. Enríquez.*<br>(Mr. Enriquez is 70 years old. He has no memory problems, but his thinking is slower than it was 5 years ago. He is retired. He drives on his own and manages the household bills. He carries out all his usual activities without help. He lives at home with his wife. They have been married for 47 years. They have 2 adult children who live nearby in their own homes. Their children visit on holidays and some weekends. Mr. Enriquez had a gene test, showing he has a medium chance (not high, not low) to develop Alzheimer's disease in the future. It is not certain he will become ill, but he has a greater chance to develop Alzheimer's disease than people whose gene test shows a low chance for Alzheimer's disease. The gene test does not show the age at which he could develop Alzheimer's disease, but his chance increases as he gets older. There is no medicine to lower Mr. Enriquez's chance of developing the disease. Mr. Enriquez and his wife tell their adult children about the gene test result. Each of their children has a high, medium, or low chance of developing Alzheimer's disease, that is distinct from Mr. Enriquez's test result.) |

Each vignette was followed by the freelisting prompt: *Enumere todas las palabras que describan la reacción del Sr. _____ al resultado de su prueba genética.* (List all the words to describe the reaction of Mr. _____ to the gene test result).
